# Amyloid and Tau Pathology are Associated with Cerebral Blood Flow in a Mixed Sample of Nondemented Older Adults with and without Vascular Risk Factors for Alzheimer’s Disease

**DOI:** 10.1101/2022.09.27.22280379

**Authors:** Cecily G. Swinford, Shannon L. Risacher, Aaron Vosmeier, Rachael Deardorff, Evgeny J. Chumin, Mario Dzemidzic, Yu-Chien Wu, Sujuan Gao, Brenna C. McDonald, Karmen K. Yoder, Frederick W. Unverzagt, Sophia Wang, Martin R. Farlow, Jared R. Brosch, David G. Clark, Liana G. Apostolova, Justin Sims, Danny J. Wang, Andrew J. Saykin

## Abstract

Alzheimer’s disease (AD) is the leading cause of dementia in individuals over 65 in the U.S. Prevalence is projected to double by 2050, but current treatments cannot stop the progression of AD. Treatments administered before severe cognitive decline may be effective; identification of biomarkers for preclinical and prodromal stages of AD is therefore imperative. Cerebral blood flow (CBF) is a potential early biomarker for AD; generally, older adults with AD have decreased CBF compared to normally aging peers. Characterization should include the relationships between CBF and AD risk factors and pathologies. We assessed the relationships between CBF quantified by arterial spin labeled MRI, hypertension, *APOE* ε4, and tau and amyloid PET in 77 older adults: cognitively normal, subjective cognitive decline, and mild cognitive impairment. Tau and amyloid aggregation were related to altered CBF, and some of these relationships were dependent on hypertension or *APOE* ε4 status. Our findings suggest a complex relationship between risk factors, AD pathologies, and CBF that warrants future studies of CBF as a potential early biomarker for AD.

## 1. Introduction

Over 6 million older adults in the U.S. have Alzheimer’s disease (AD)^1^, and this number is projected to double over the next thirty years (Alzheimer’s Disease Facts and Figures, 2021). The results of multiple studies and clinical trials suggest that an effective treatment for AD may need to be preventative (Jessen et al., 2014; Crous-Bou et al., 2017). Excluding at least one promising exception (van Dyck et al., 2023), treatments that target amyloid beta (Aβ) and/or tau pathology have generally been unable to restore cognitive function after cognitive decline has occurred (Lu et al., 2020; Congdon and Sigurdsson, 2018). Preventative treatment will require reliable biomarkers for the preclinical stages of AD so that individuals most at risk can be accurately identified. One potential early biomarker that warrants further characterization is quantitative cerebral blood flow (CBF).

CBF is known to be decreased in individuals with AD compared to healthy individuals of the same age (Alsop et al., 2000; Swinford et al., 2022; Zhang et al., 2021), independently of brain atrophy (Binnewijzend et al., 2013). Additionally, vascular risk factors including hypertension, diabetes mellitus, and obesity are risk factors for AD (Gorelick, 2004), and they have been shown to have an effect on CBF (Bangen et al., 2014). Chronic hypertension, in particular, has been shown to cause cerebral microvascular damage, which both worsens and is worsened by Aβ aggregation (Iadecola and Gottesman, 2019). It has been suggested that altered CBF may occur very early in the course of AD, before cognitive decline and potentially even prior to Aβ and tau aggregation (Iadecola, 2004).

However, the spatial and temporal patterns of CBF alteration over the course of AD have not been fully characterized. It has been shown that CBF increases in some brain regions, especially in preclinical and prodromal AD, known as subjective cognitive decline (SCD) and mild cognitive impairment (MCI; Beishon et al., 2017; Dai et al., 2009). However, most individuals have decreased CBF by the time they meet criteria for AD (Swinford et al., 2022; Wierenga et al., 2014). Increased CBF in SCD and early MCI may compensate to maintain a level of cognitive functioning despite pathologies such as Aβ and/or tau aggregation for a limited time. It is important to elucidate the spatial and temporal relationships between altered CBF, other AD-related pathologies, and vascular risk factors. Here, in older adults who are cognitively normal (CN), SCD, and MCI, we assessed the relationships between CBF and global cortical Aβ and medial temporal lobe (MTL) tau pathology using arterial spin labeled (ASL) magnetic resonance imaging (MRI) and positron emission tomography (PET). We also assessed the interaction effects of hypertension and *APOE* ε4 positivity on these relationships. We hypothesized that Aβ and tau pathologies would negatively correlate with CBF overall, although localized regions of positive correlation would be present as well, given the early stage of disease in this sample. We also hypothesized that individuals with hypertension or at least one *APOE* ε4 allele would be less likely to have a compensatory positive correlation between CBF and AD-related pathologies.

## 2. Methods

### 2.1 Participants

This study included 77 participants (30 CN, 24 SCD, 23 MCI) from the Indiana Alzheimer’s Disease Research Center who were at least sixty years of age. All participants completed T1-weighted structural and pseudocontinuous arterial spin labeled (pCASL) MRI scans, Aβ PET scan with either [^18^F]florbetapir or [^18^F]florbetaben, and tau PET scan with [^18^F]flortaucipir. All participants also had a clinical assessment including administration of the objective neuropsychological test battery Uniform Data Set version 3 (UDS 3.0; Besser et al., 2018) and *APOE* genotyping. Diagnoses were determined by clinician consensus. SCD was defined as a score of 20 or more on the first 12 items of the 20-item Cognitive Change Index (CCI-20; Saykin et al., 2006), reflecting increased subjective memory concerns, with or without increased levels of informant-based concerns (Jessen et al., 2014), and the absence of a measurable cognitive deficit on the UDS 3.0. Using standard criteria, an individual was diagnosed with MCI if the participant and/or an informant had a significant complaint about their cognition, and the participant scored 1.5 standard deviations or more below age-, education-, and sex-adjusted normative data on objective tests of cognitive functioning in the UDS 3.0 either in memory or another cognitive domain. Additionally, individuals diagnosed with MCI, by definition, do not show a significant decline in daily functioning.

Hypertension was defined as a previous diagnosis of hypertension via self-report near the time of imaging. It has been shown that midlife hypertension, rather than hypertension later in life, confers the greatest risk for AD later in life (Lennon et al., 2021; Whitmer et al., 2005). Some hypertensive individuals were taking antihypertensive medication at the time of the study. A vascular risk score was calculated for each participant. Each participant received one vascular risk point for each of the following conditions: reported diabetes and/or use of diabetes medication, reported hypercholesterolemia and/or use of medication for high cholesterol, obesity as defined by BMI greater than 30, and blood pressure that meets criteria for hypertension (systolic blood pressure equal to or greater than 130 and/or diastolic blood pressure equal to or greater than 80) taken from the appointment closest to the ASL MRI scan. Vascular risk scores ranged from 0 to 4 points. All procedures were approved by the Indiana University School of Medicine Institutional Review Board. Informed consent was obtained according to the Declaration of Helsinki and the Belmont Report.

### 2.2 pCASL and T1-weighted MRI

All MRI scans were acquired on the same Siemens 3T Prisma scanner with a 64-channel head/neck radiofrequency coil. After an initial T1-weighted structural scan (sagittal 3D MPRAGE) each participant completed a 3D pseudo-continuous ASL (pCASL) MRI scan.

Processing steps were performed using Statistical Parametric Mapping 12 (SPM12; Wellcome Department of Cognitive Neuroscience, London, UK) and Matlab R2022a as previously described (Butcher et al., 2021; Wang et al., 2003). To calculate CBF from pCASL images, pairs of pCASL images were subtracted and then averaged to obtain a mean perfusion image. A standard single-compartment perfusion model (Alsop et al., 2015) was used to calculate mean CBF maps from the mean perfusion images (http://cfn.upenn.edu/perfusion/software.htm). Using each individual’s gray matter tissue map from the structural MRI, an individualized gray matter mask was applied to each mean CBF map, and the cerebellum was excluded in order to remove individual variability in this region, which is common with pCASL imaging. CBF was extracted from the resulting masked CBF map from frontal, parietal, temporal, and occipital lobes and limbic regions of interest (ROIs) using Automated Anatomical Labeling (AAL) version 1 and the MarsBaR toolbox (Brett et al., 2002).

### 2.3 Aβ PET

Each participant underwent either a [^18^F]florbetapir (AmyVid; Eli Lilly and Co.) or a [^18^F]florbetaben (Neuraceq; Life Molecular Imaging, formerly Piramal Imaging) scan. Aβ PET data was recorded in four 5-minute frames and reconstructed using parameters defined in the Alzheimer’s Disease Neuroimaging Initiative (ADNI) 2 protocol (Jagust et al., 2015; https://adni.loni.usc.edu/methods/pet-analysis-method/pet-analysis/). Each 5-minute frame was spatially aligned to the participant’s T1-weighted structural MRI using SPM12. The spatially aligned PET images were averaged to create a single image from the total acquisition time period for each individual, and standardized uptake value ratio (SUVR) images were generated by intensity normalizing with the average radioactivity from a whole cerebellar region of interest from the Centiloid project (http://www.gaain.org/Centiloid-project/). For this study, we used the global cortical Aβ measure calculated in Centiloid units, a unified metric that allowed us to directly compare data from both scan types. The global cortical ROI was determined by the Centiloid project and is the ROI for which use of Centiloid units is intended. Both types of scans were processed with the Centiloid algorithm at a voxel-wise level as defined by the Centiloid project (Risacher et al., 2021). Global cortical Centiloid data were then extracted from both scan types using the MarsBaR toolbox (Brett et al., 2002).

### 2.4 Tau PET

All participants underwent a [^18^F]flortaucipir PET scan. Individuals were imaged on the same Siemens Biograph mCT PET/CT scanner for 30 minutes; data were binned into six 5-minute frames, and scans were reconstructed using a standard scanner software program (Siemens, Knoxville, TN) according to the ADNI2 protocol (Jagust et al., 2015; https://adni.loni.usc.edu/methods/pet-analysis-method/pet-analysis/). The 5-minute frames were spatially aligned to each participant’s T1 image and normalized to MNI space, averaged to create a single image, intensity normalized by the cerebellar crus (Schwarz et. al., 2016) to create SUVR images, and smoothed, all using SPM12. For this study, we extracted the MTL SUVR. This region was chosen because it is the main location of tau aggregation in early stages of AD, including SCD and MCI, so it was considered the region most likely to correlate with disease severity in our sample. The constituent ROIs were generated from FreeSurfer 6.0 (Fischl and Dale, 2000; http://surfer.nmr.mgh.harvard.edu/), and the bilateral mean MTL (average of fusiform gyri, parahippocampal gyri, and entorhinal cortex) SUVR values were extracted for each participant using MarsBaR.

### 2.5 Statistical Analyses

The demographic and clinical variables, as well as global CBF, total gray matter volume, and Aβ and tau PET measures were compared between diagnostic groups using one-way ANOVA for continuous variables and chi-square test for categorical variables. Total gray matter volume was included as a covariate in subsequent analyses to confirm that decreased CBF is indicative of hypoperfusion rather than a decreased need for CBF in line with decreased brain tissue. Stepwise multiple regression models (entry p=0.10, removal p=0.20) with dependent variable regional CBF and independent variables, age, sex, diagnostic group, total gray matter volume (from T1-weighted image gray matter estimates by SPM12 tissue volume utility), reported hypertension status, *APOE* ε4 status, vascular risk score, and use of antihypertensive medication were used to determine demographic and clinical variables that correlated with CBF in this sample. Additionally, Aβ and tau PET metrics were treated as the dependent variables in multiple regression models with age, sex, diagnostic group, total gray matter volume, reported hypertension status, *APOE* ε4 status, vascular risk score, and use of antihypertensive medication as independent variables to assess correlations between Aβ and tau PET and demographic and clinical variables. Partial Pearson correlations assessed the relationships between regional CBF and Aβ or tau PET metrics, with age, sex, diagnostic group, total gray matter volume as covariates. Reported hypertension status, *APOE* ε4 status, vascular risk score, and use of antihypertensive medication were added as covariates. Voxel-wise analyses using multiple regression with CBF maps as the dependent variable, Aβ or tau PET metric as the independent variable, and age, sex, diagnostic group and total gray matter volume as covariates were used to explore associations that may not have been detectable when using the lobar regions of interest. Voxel-wise analyses were also used to explore the effects of the interactions between Aβ or tau PET metrics and reported hypertension or *APOE* ε4 status on CBF with Aβ or tau PET metric, reported hypertension or *APOE* ε4 status, age, sex, diagnostic group and total gray matter volume as covariates. CBF was extracted from significant clusters and modeled as the dependent variable in linear regression to visualize interactions. Results are listed at threshold *p*≤0.001 and cluster size (*k*)≥50 voxels (uncorrected) for clusters with peak voxels within labeled gray matter regions. All voxel-wise analyses were completed using SPM12. Other statistical analyses and all graphs were created in IBM SPSS 25.

## 3. Results

### 3.1 Demographics

Demographics and clinical variables are given in Table 1. The MCI group had a greater global cortical Centiloid value than the CN (Bonferroni *T*=3.54, *p*=0.002) and SCD (Bonferroni *T*=3.08, *p*=0.009) groups (F(2, 76) = 7.24, *p*=0.001). Diagnostic groups also differed by prevalence of self-reported hypertension (*χ*^2^ (2)=11.09, *p*=0.004) but did not differ by blood pressure taken at the time of scan (*p*>0.05). The SCD group had the highest prevalence of reported hypertension, followed by MCI, and then by CN. There is a trend level difference in prevalence of antihypertensive medication use (*χ*^2^ (2)=5.59, *p*=0.061). Prevalence was highest in the SCD group, followed by MCI and then CN. Neither global CBF nor the vascular risk score differed by diagnostic group (*p*>0.05).

**Table 1.** Demographic and clinical variables by diagnostic group.

|  | All (n=77) | CN (n=30) | SCD (n=24) | MCI (n=23) | Group Comparison *p* |
| --- | --- | --- | --- | --- | --- |
| Age | 71.69 ± 6.75 | 71.03 ± 6.66 | 71.04 ± 6.88 | 73.22 ± 6.78 | 0.436 |
| Sex (F) | 64.9% | 76.7% | 66.7% | 47.8% | 0.091 |
| Education | 16.44 ± 2.78 | 16.73 ± 2.55 | 16.54 ± 2.80 | 15.96 ± 3.11 | 0.595 |
| Race | 14.3% | 16.7% | 20.8% | 4.3% | 0.242 |
| Ethnicity | 1.3% | 0% | 0% | 4.3% | 0.304 |
| GM Volume (mm<sup>3</sup>) | 571,221 ± 58,254 | 573,685 ± 58,882 | 574,368 ± 58,392 | 564,723 ± 59,372 | 0.819 |
| Reported Hypertension | 51.9% | 30.0% | 75.0% | 56.5% | 0.004*<br>SCD > MCI > CN |
| Global CBF APOE ε4 positive | 42.9% | 43.3% | 33.3% | 52.2% | 0.426 |
| Amyloid (Aβ) PET (Centiloid) | 19.79 ± 32.17 | 10.31 ± 24.43 | 12.70 ± 26.74 | 39.54 ± 38.16 | 0.001*<br>MCI > CN&SCD |
| Tau PET (SUVR) | 1.11 ± 0.11 | 1.10 ± 0.09 | 1.09 ± 0.06 | 1.15 ± 0.16 | 0.174 |
| Vascular Risk Score | 1.78 ± 1.02 | 1.60 ± 1.13 | 1.87 ± 1.12 | 1.91 ± 0.73 | 0.471 |
| Systolic BP | 134.31 ± 19.00 | 134.53 ± 21.56 | 130.75 ± 13.51 | 137.74 ± 20.41 | 0.456 |
| Diastolic BP | 71.73 ± 9.46 | 72.00 ± 9.23 | 71.46 ± 10.67 | 71.65 ± 8.81 | 0.978 |
| Hypertension | 61.0% | 60.0% | 58.3% | 65.2% | 0.880 |
| BMI | 28.25 ± 5.26 | 27.61 ± 5.36 | 29.15 ± 5.87 | 28.15 ± 4.50 | 0.568 |
| Reported Diabetes | 18.2% | 10.0% | 29.2% | 17.3% | 0.191 |
| Reported Hypercholesterolemia | 66.2% | 60.0% | 62.5% | 78.3% | 0.340 |
| HT Med | 53.2% | 36.7% | 66.7% | 60.9% | 0.061^ |
| HLD Med | 54.5% | 53.3% | 45.8% | 65.2% | 0.405 |
| DM Med | 15.6% | 13.3% | 25.0% | 8.7% | 0.278 |

PET: positron emission tomography, CN: cognitively normal, SCD: subjective cognitive decline, MCI: mild cognitive impairment, GM: gray matter, APOE: apolipoprotein E, CBF: cerebral blood flow, SUVR: standardized uptake value ratio

### 3.2 CBF vs. Demographic and Clinical Variables

Age and reported hypertension were negatively correlated with CBF in all lobes, and female sex was positively correlated with CBF in limbic and occipital lobes (Table 2). When this analysis was repeated with stratification by diagnostic group, age was negatively correlated with CBF in the CN group, reported hypertension was negatively correlated with CBF in the frontal lobe in the MCI group, and female sex was positively correlated with CBF in the MCI group (Supplementary Tables 1 and 2). Voxel-wise regressions were used to spatially model these relationships, revealing more widespread negative correlations between reported hypertension and CBF in the MCI group when the vascular risk score and use of antihypertensive medication were included as covariates (Figures 1 and 2, Supplementary Figures 1 and 2, Supplementary Tables 3-7).

**Table 2.** Correlation of age, reported hypertension status, and female sex with lobar CBF.

| | Independent Variable retained in Model | Standardized Coefficient ($\beta$) | *t* | *p* |
| --- | --- | --- | --- | --- |
| Frontal | Age | −0.343 | −3.362 | 0.001 |
|  | Reported Hypertension | −0.315 | −3.095 | 0.003 |
| Limbic | Age | −0.307 | −2.947 | 0.004 |
|  | Reported Hypertension | −0.210 | −2.009 | 0.048 |
|  | Sex | 0.219 | 2.104 | 0.039 |
| Parietal | Age | −0.307 | −2.901 | 0.005 |
|  | Reported Hypertension | −0.262 | −2.478 | 0.015 |
| Temporal | Age | −0.334 | −3.194 | 0.002 |
|  | Reported Hypertension | −0.267 | −2.555 | 0.013 |
| Occipital | Age | −0.298 | −2.924 | 0.005 |
|  | Reported Hypertension | −0.216 | −2.114 | 0.038 |
|  | Sex | 0.278 | 2.724 | 0.008 |

**Figure 1.**
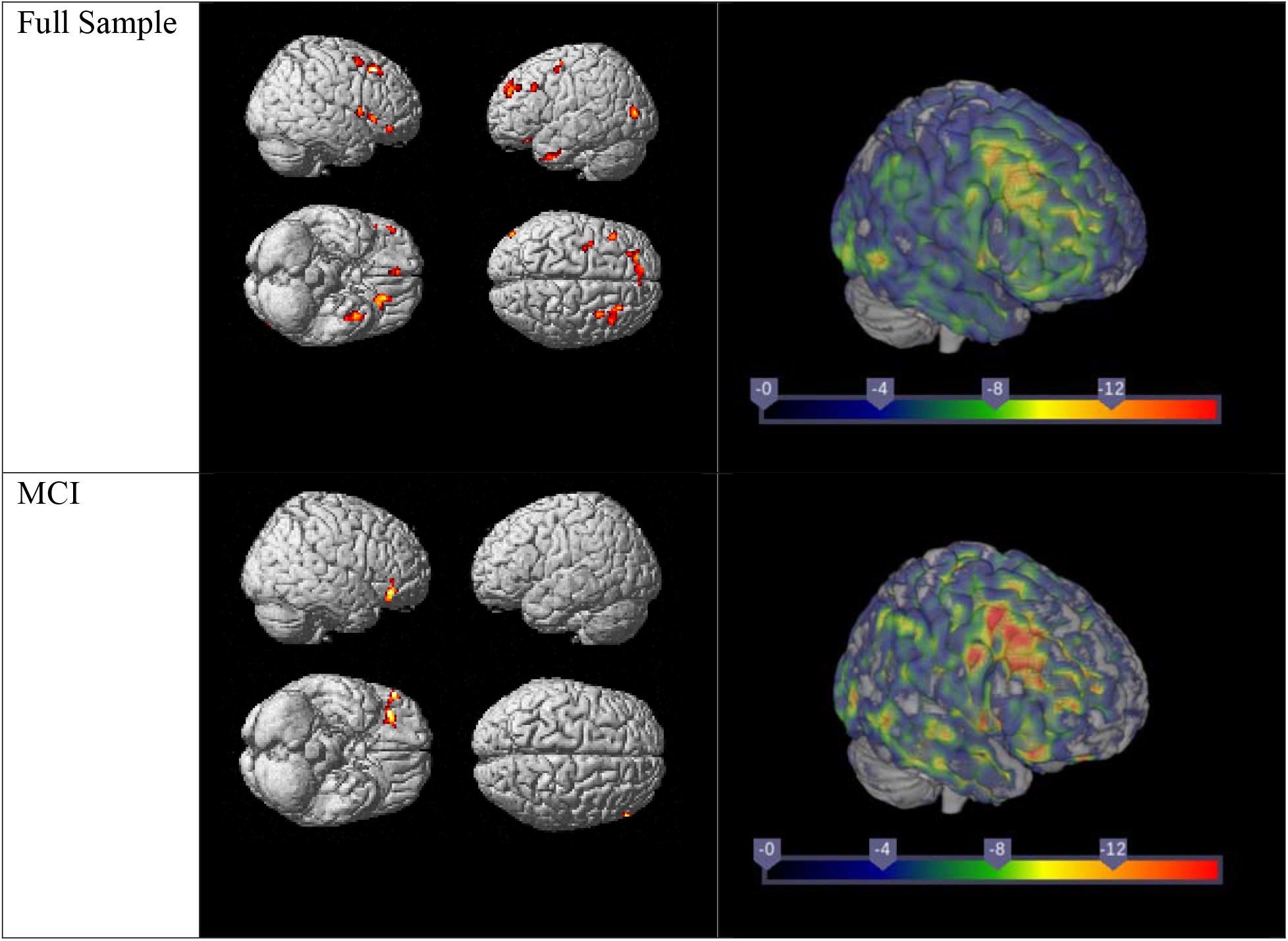
Negative correlations between voxel-wise CBF and reported hypertension status. Covariates are age, sex, diagnostic group and total gray matter volume. Images in left column illustrated at *p* 0.001; *k* 50. Right column shows maps.

**Figure 2.**
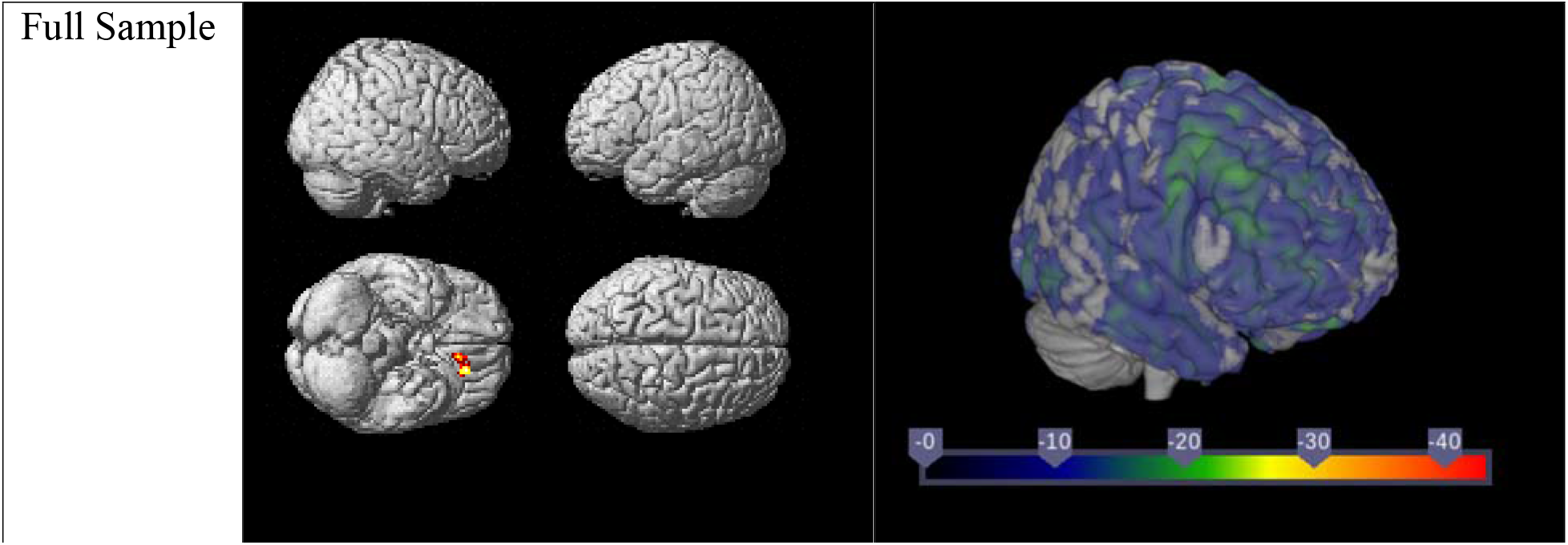
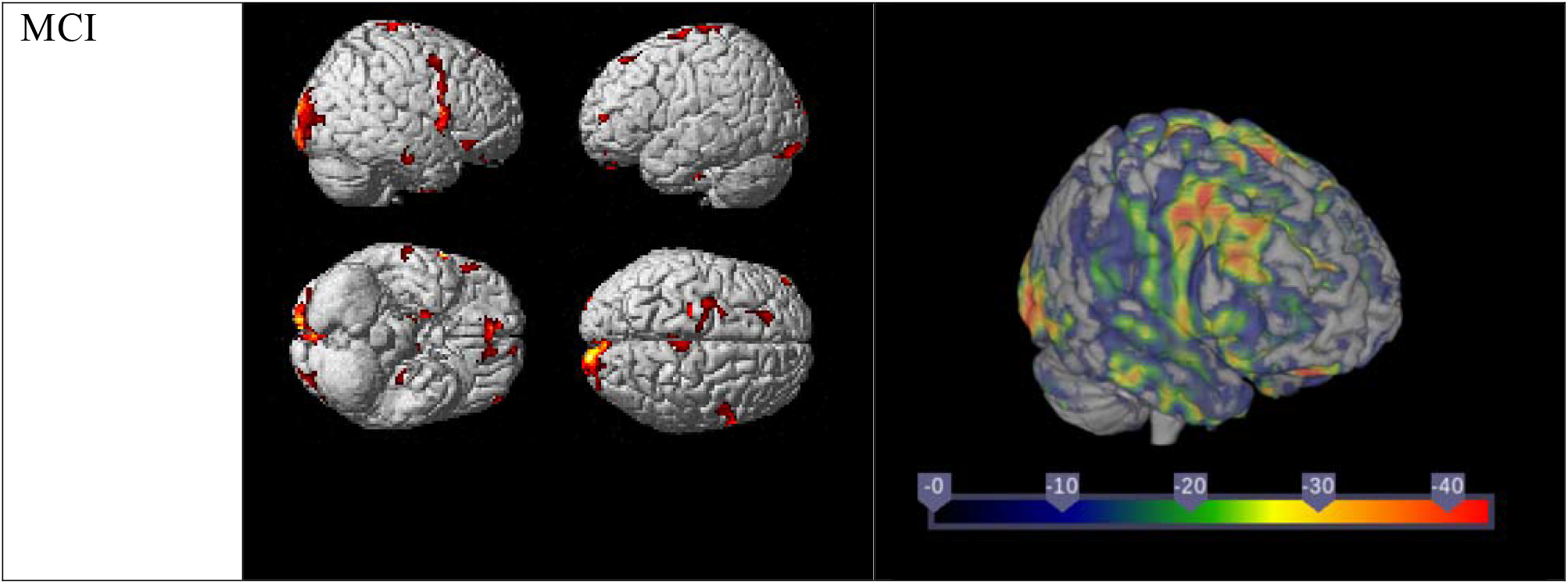
Negative correlations between voxel-wise CBF and reported hypertension status. Covariates are age, sex, diagnostic group, total gray matter volume vascular risk score and use of antihypertensive medication. Images in left column illustrated at *p* 0.001; *k* 50. Right column shows maps.

### 3.3 Tau and Aβ PET vs. Demographic and Clinical Variables

The global cortical Centiloid value was positively associated with *APOE* ε4 status (β=0.388, *t*=3.992, *p*<0.001) and MCI (β=0.356, *t*=3.657, *p*<0.001). None of the independent variables were entered into the model for MTL tau SUVR (*p*>0.05). When the analyses were repeated with stratification by diagnostic group, the global cortical Centiloid value was positively associated with *APOE* ε4 status in the CN group (β=0.455, *t*=2.703, *p*=0.012) and in the MCI group (β=0.476, *t*=2.483, *p*=0.022). The MTL tau SUVR was positively associated with age in the SCD group (β=0.517, *t*=2.836, *p*=0.010) and negatively associated with age (β=-0.369, *t*=-2.103, *p*=0.048) and reported hypertension (β=-0.545, *t*=-3.104, *p*=0.006) in the MCI group.

### 3.4 Tau PET vs. Regional CBF

Partial correlations between lobar CBF and MTL tau SUVR were not significant regardless of whether reported hypertension, *APOE* ε4 status, vascular risk score, and use of antihypertensive medication were included as covariates (*p*>0.05). There was a trend level negative correlation between MTL tau SUVR and parietal lobe CBF in the SCD group (*r*=-0.416, *p*=0.061) with age, sex, and total gray matter volume as covariates. Using voxel-wise regression models with age, sex, diagnostic group and total gray matter volume as covariates, MTL tau SUVR was negatively correlated with CBF in the left parahippocampal gyrus in the full sample (*p*≤0.001, *k*≥50; Figure 3, Supplementary Table 8). MTL tau SUVR was negatively correlated with CBF in the left primary sensory, right primary motor, and right visual motor regions in the SCD group (*p*≤0.001, *k*≥50; Figure 3, Supplementary Table 9).

**Figure 3.**
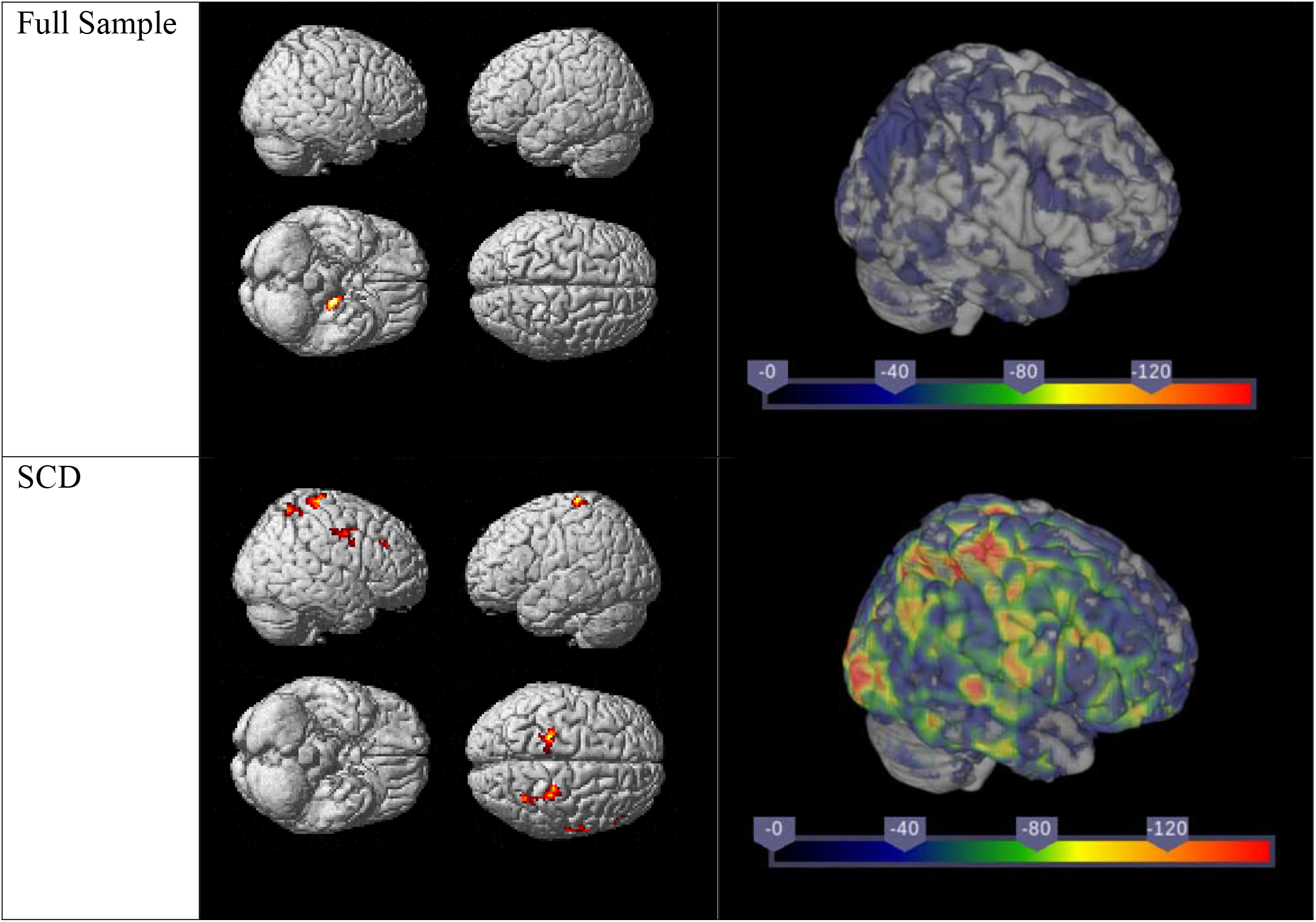
Negative correlations between voxel-wise CBF and MTL tau SUVR. Covariates are age, sex, diagnostic group, and total gray matter volume. Images in left column illustrated at *p* 0.001; *k* 50. Right column shows maps.

There were positive correlations between the MTL tau SUVR and reported hypertension status interaction variable and CBF in several brain regions in the SCD group (*p*≤0.001, *k*≥50; Supplementary Table 10). There were negative correlations between the MTL tau SUVR and *APOE* ε4 status interaction variable and spatially widespread CBF in the SCD group (*p*≤0.001, *k*≥50; Figure 4, Supplementary Table 11) and a positive correlation between the MTL tau SUVR and *APOE* ε4 status interaction variable and CBF in the right fusiform in the MCI group (*p*≤0.001, *k*≥50; Supplementary Table 12).

**Figure 4.**
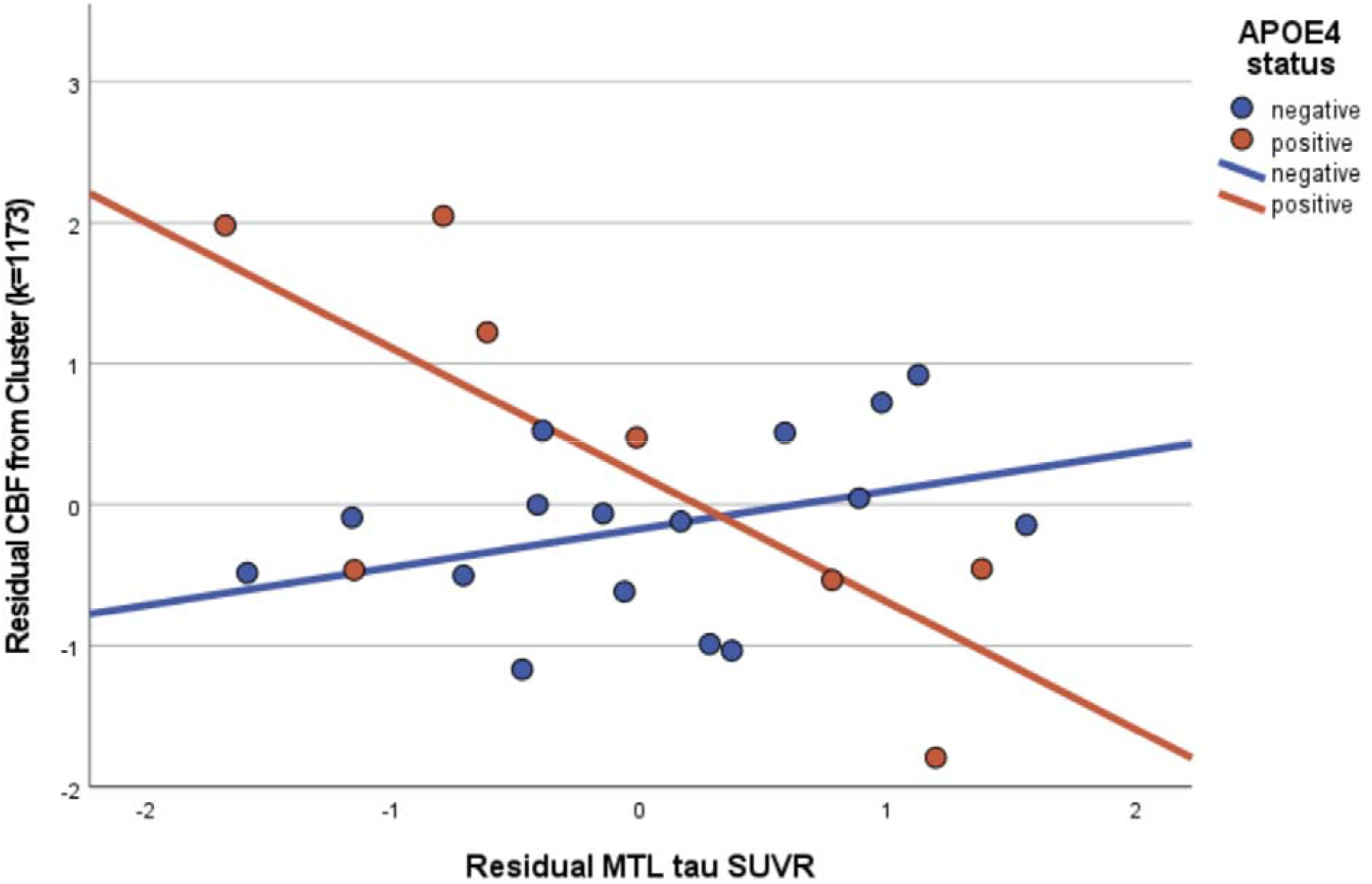
Scatterplot of the positive correlation between voxel-wise CBF and MTL tau SUVR in *APOE* _ε_4 negative individuals and the negative correlation between voxel-wise CBF and MTL tau SUVR in *APOE* _ε_4 positive individuals in the SCD group. Both the CBF and MTL tau SUVR plotted are standardized residuals from regressions with age, sex, and total gray matter volume. *APOE* _ε_4 negative R^2^=0.137; *APOE* _ε_4 positive R^2^=0.559.

### 3.5 Aβ PET vs. Regional CBF

Partial correlations between lobar CBF and global cortical Centiloid were not significant regardless of whether reported hypertension, *APOE* ε4 status, vascular risk score, and use of antihypertensive medication were included as covariates (*p*>0.05). Using voxel-wise regression models with age, sex, diagnostic group and total gray matter volume as covariates, there were no statistically significant correlations between CBF and global cortical Centiloid at the *p*≤0.001, *k*≥50 level. However, there was a positive correlation between global cortical Centiloid and CBF in the right visual association region in the CN group (*p*≤0.001, *k*≥50; Figure 5, Supplementary Table 13) and a negative correlation between global cortical Centiloid and CBF in the right fusiform in the MCI group (*p*≤0.001, *k*≥50; Figure 5, Supplementary Table 14). There was a positive correlation between the global cortical Centiloid and reported hypertension interaction variable and CBF in the left visual motor region in the whole sample (*p*≤0.001, *k*≥50; Figure 6, Supplementary Table 15). There was a positive correlation between the global cortical Centiloid and reported hypertension status interaction variable and CBF in the right hippocampus in the SCD group (*p*≤0.001, *k*≥50; Supplementary Table 16) and a positive correlation between the global cortical Centiloid and reported hypertension status interaction variable and CBF in the left frontal eye fields in the MCI group (*p*≤0.001, *k*≥50; Supplementary Table 17).

**Figure 5.**
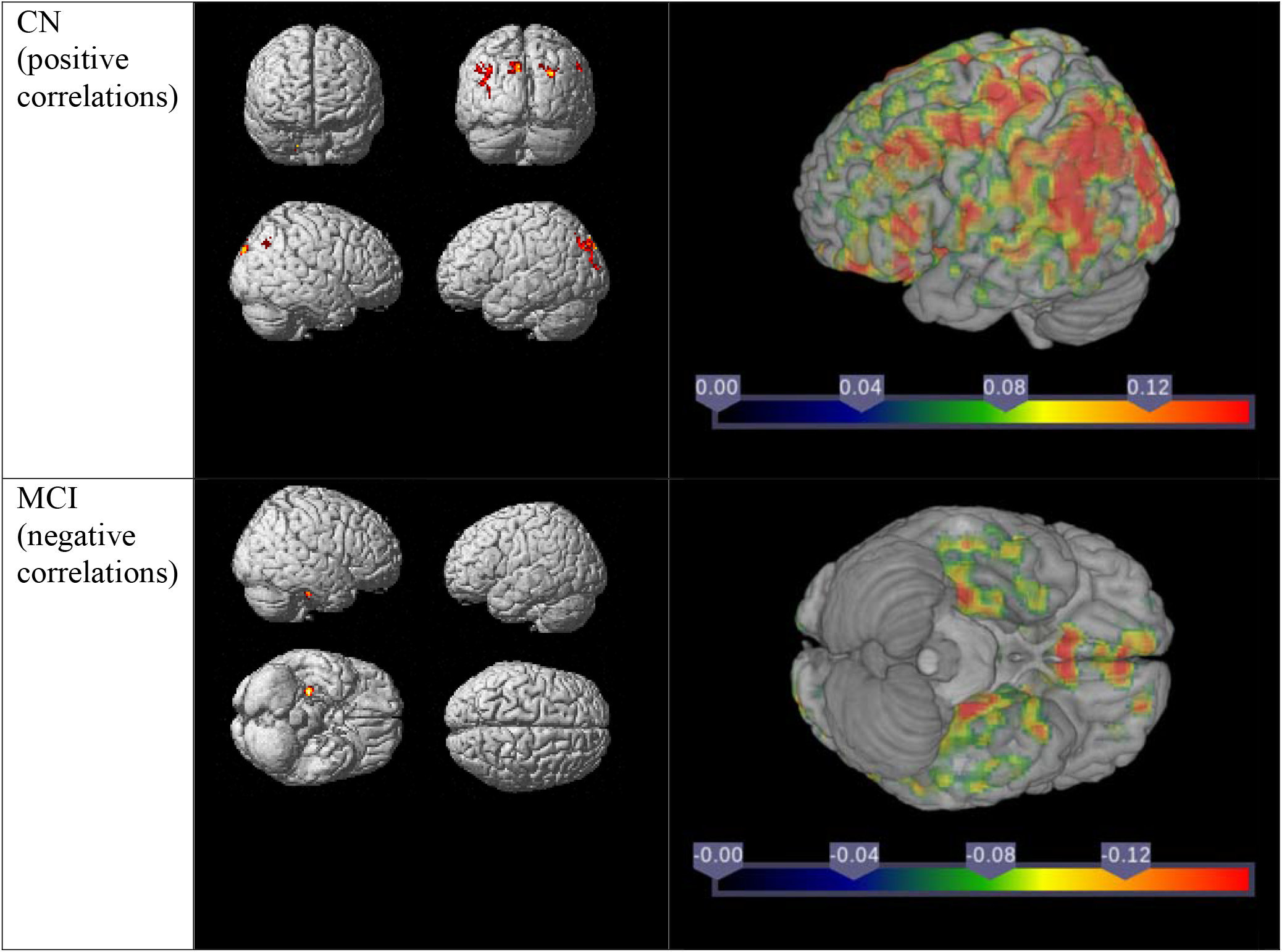
Correlations between voxel-wise CBF and global cortical Centiloid. Covariates are age, sex, and total gray matter volume. Images in left column illustrated at *p* 0.001; *k* 50. Right column shows maps.

**Figure 6.**
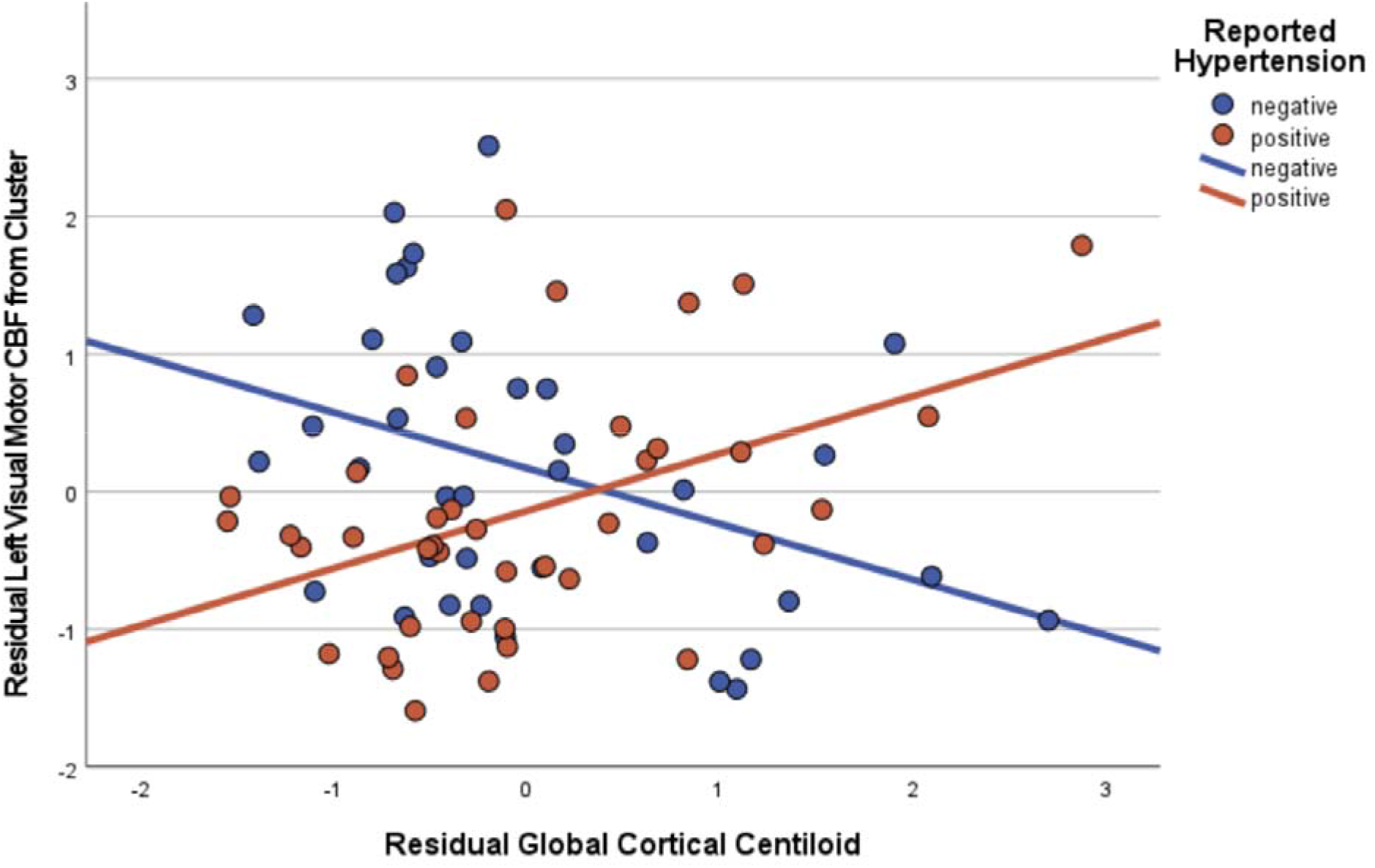
Scatterplot of the negative correlation between voxel-wise CBF and global cortical Centiloid in individuals without reported hypertension (negative) and the positive correlation between voxel-wise CBF and global cortical Centiloid in individuals with reported hypertension (positive) in the whole sample. Both the CBF and global cortical Centiloid plotted are standardized residuals from regressions with age, sex, and total gray matter volume. Reported hypertension negative R^2^=0.155; reported hypertension positive R^2^=0.196.

## 4. Discussion

We found associations between CBF, global cortical Centiloid, MTL tau SUVR, and vascular risk factors in this sample of CN, SCD, and MCI older adults. Both reported hypertension and MTL tau SUVR were negatively associated with CBF among and within diagnostic groups in exploratory voxel-wise analyses. Although altered, some negative associations between reported hypertension and CBF remained when vascular risk score and use of antihypertensive medication were taken into account. Global cortical Centiloid was positively associated with CBF in the CN and negatively associated with CBF in the MCI group in voxel-wise analyses. Finally, there were effects of AD pathology and vascular risk factor interaction variables on CBF in voxel-wise analyses. These include a negative effect of the MTL tau SUVR and *APOE* ε4 status interaction variable on CBF in the SCD group and a positive effect of the global cortical centiloid and reported hypertension status interaction variable on CBF in the full sample. While these findings generally follow our hypotheses, they also suggest that the relationships among these factors are complex and dynamic.

Previous studies, briefly described below, concerning the relationship between tau and CBF have reported negative correlations that track both spatially and temporally with disease severity and brain dysfunction (Ashford et al., 2000; Gomez-Isla et al., 1997). Animal models of hypoperfusion develop increased tau pathology (Raz et al., 2019; Zhang et al., 2010), and animal models of tau pathology develop hypoperfusion (Bennett et al., 2018; Jaworski et al., 2011; Lourenco et al., 2017; Park et al., 2020). In human studies in preclinical and/or clinical AD, there have been findings of negative correlations between tau pathology and CBF in temporal, parietal, and occipital lobes, and entorhinal cortex (Albrecht et al., 2020; Rubinski et al., 2021; Visser et al., 2020). Studies in CN individuals report a mix of results; Hays et al. (2020) reported a negative correlation between CSF tau and CBF in the anterior cingulate cortex, while Stomrud et al. (2012) reported a positive correlation between tau and CBF in the frontotemporal border zone and a negative correlation between tau and CBF in the medial frontal lobe. Another study reported that asymptomatic individuals with the same amount of tau pathology as individuals with MCI had increased CBF in the MTL and thalamus over time (Codispoti et al., 2012). Although there are fewer studies focused on the relationship between tau pathology and CBF compared to Aβ pathology, it appears that a similar overall pattern of findings is emerging, with some positive correlations in early disease stages and negative correlations in later stages.

Negative correlations between Aβ pathology and CBF have been heavily documented. For example, several animal models of chronic hypoperfusion have been shown to develop Aβ pathology (Chan et al., 2018; de la Torre et al., 2003; Faraco et al., 2016; Khan et al., 2015; Liang et al., 2015), and animal models of Aβ pathology, likewise, develop decreased CBF (Klohs et al., 2014; Niwa et al., 2001). Studies involving participants with preclinical and/or clinical AD have shown negative correlations between Aβ pathology and CBF in entorhinal regions, the precuneus, and throughout the cortex (Mattsson et al., 2014; Rodell et al., 2016). Chen et al. (2015), on the other hand, reported a positive correlation between Aβ and CBF in the anterior cingulate gyrus in a mixed sample including AD patients. Michels et al. (2016) and Tosun et al. (2016) both reported negative correlations between Aβ PET and CBF in regions including frontal, temporal, parietal, and occipital lobes, cingulate cortex, and cerebellum in disease stages prior to AD. In a sample of Aβ -positive CN individuals, Aβ was positively correlated with CBF in frontal and temporal lobes, insula, dorsal striatum, hippocampus, and amygdala (Fazlollahi et al., 2020).

Interacting relationships between vascular risk factors and AD pathologies and symptoms have been previously studied. Specifically, two papers (Wang et al., 2019; Zlatar et al., 2016) reported a positive correlation between CBF and cognition in *APOE* ε4-negative CN individuals and a negative correlation between CBF and cognition in *APOE* ε4-positive CN individuals. Hays et al. (2020) found a negative relationship between tau in the cerebrospinal fluid and CBF in the anterior cingulate in *APOE* ε4-positive but not *APOE* ε4-negative individuals. *APOE* ε4 is associated with increased Aβ aggregation, most likely by reducing the clearance of Aβ from the brain and increasing its production (Castellano et al., 2011; Donahue and Johanson, 2008; Mahley et al., 2006; Wahrle et al., 2007). It has also been reported that *APOE* ε4 carriers have increased levels of tau in the MTL (Jack et al., 2020; Ossenkoppele et al., 2016).

Our findings are generally in line with current literature. As we expected, both reported hypertension and MTL tau SUVR are negatively correlated with CBF in this sample. Reported hypertension appeared to be particularly associated with decreased CBF in the MCI group, while MTL tau SUVR appeared to be particularly associated with decreased CBF in the SCD group. In the SCD group, regions of CBF were negatively correlated with MTL tau SUVR in *APOE* ε4 positive individuals and positively correlated with MTL tau SUVR in *APOE* ε4 negative individuals. It is possible that individuals with *APOE* ε4 are more susceptible to hypoperfusion related to tau pathology in early disease stages. Global cortical Centiloid was positively correlated with CBF in the CN group but negatively correlated with CBF in the MCI group. This follows the general trend of compensatory mechanisms in early disease stages followed by loss of compensation and deterioration of brain function as the disease progresses. Our finding of a positive correlation between global cortical Centiloid and CBF in individuals with reported hypertension but a negative correlation between global cortical Centiloid and CBF in individuals without reported hypertension across all diagnostic groups suggests that Aβ aggregation and hypertension may not have strictly additive effects on brain function. This interaction was not in the direction that we expected, but there is not yet a standard time course for compensation versus failure of compensation, and it likely differs for each individual based on comorbidities, cardiovascular health, lifestyle choices like diet and exercise, and demographic, genetic, and environmental variables. Our findings suggest that the combined effects of vascular risk factors, AD pathology, and AD-specific risk factors on CBF warrant further research with large and diverse patient samples.

This study was limited by its modest sample size. Some regions of association between voxel-wise CBF and PET biomarkers did not reach statistical significance, and a larger sample size could have led to more statistically significant results suggesting a clearer picture of correlative patterns. Future studies should use larger sample sizes of individuals with preclinical AD so that statistical power is adequate to elucidate the complex relationships between CBF, Aβ, tau, and other AD-related pathologies. Our sample was also homogenous in terms of ethnoracial and educational backgrounds, making our findings most applicable to a subset of the population of older adults at risk for AD. It will be important to utilize diverse samples in future studies. Additionally, our use of MTL tau SUVR, while it is the region most likely involved in early stage tau aggregation did not allow us to take atypical patterns of tau aggregation into account in this study. Finally, we defined hypertension by self-report rather than by a threshold blood pressure at the time of scanning, and some hypertensive individuals were taking antihypertensive medications. We included blood pressure as part of our vascular risk score and included this score and use of antihypertensives in a number of our models, but future studies with a narrower definition of hypertension may be beneficial.

### 4.1 Conclusions

Our findings suggest complex spatial and temporal relationships among genetic and vascular risk factors, AD pathologies, and CBF in older adults at risk for AD. A reliable early biomarker for AD would allow us to identify those at risk for AD so that they can enter clinical trials and/or receive treatment, track disease progression in both treated and untreated individuals, and perhaps help elucidate the complex mechanisms that initiate AD and drive its progression. Future studies assessing the potential of CBF as an early biomarker for AD are warranted.

## Data Availability

All data produced in the present study are available upon reasonable request to the authors.

## ^1^ Abbreviations

Aβ: amyloid beta
AAL: Automated Anatomical Labeling
AD: Alzheimer’s disease
ADNI: Alzheimer’s Disease Neuroimaging Initiative
APOE: apolipoprotein E
ANCOVA: analysis of covariance
ANOVA: analysis of variance
ASL: arterial spin labeled
CBF: cerebral blood flow
CCI: Cognitive Change Index
CN: cognitively normal
CSF: cerebrospinal fluid
FWE: Family-Wise Error
FWHM: full width at half maximum
MCI: mild cognitive impairment
MNI: Montreal Neurological Institute
MRI: magnetic resonance imaging
MTL: medial temporal lobe
pCASL: psuedocontinuous arterial spin labeled
PET: positron emission tomography
PIB: Pittsburgh compound B
ROI: region of interest
SCD: subjective cognitive decline
SPM: Statistical Parametric Mapping
SPSS: Statistical Package for the Social Sciences
SUVR: standardized uptake value ratio

## Acknowledgements

John D. West, MSEng, Indiana University School of Medicine, played a primary role in optimizing and automating the pCASL processing pipeline, and the authors are grateful for his contributions to this research which we dedicate to his memory. We are grateful to the research participants and their families without whose generosity and commitment this study would not have been possible.

This study was funded in part by the following grants from the National Institute on Aging: K01 AG049050, R01 AG061788, P30 AG010133, P30 AG072976, R01 AG019771, R01 AG057739, U01 AG024904, R01 AG068193, T32 AG071444, U01 AG068057, and U01 AG072177 and the National Library of Medicine: R01 LM013463. In kind support for tau PET tracer precursor was provided by Avid Radiopharmaceuticals, a subsidiary of Eli Lilly and Company

