## Supplemental File for "Amyloid and Tau Pathology are Associated with Cerebral Blood Flow in a Mixed Sample of Nondemented Older Adults with and without Vascular Risk Factors for Alzheimer’s Disease"

Supplementary Methods

2.2 pCASL and T1-weighted MRI

The full MRI protocol consisted of MPRAGE, FLAIR, high resolution hippocampus imaging, DTI, fMRI, HYDI DTI, pCASL, and finally SWI over a total of roughly 58 minutes. The MPRAGE scan had the following parameters: repetition time (TR) 2300 ms, echo time (TE) 2.95 ms, field of view 270 mm, 176 slices, and voxel size 1.1 x 1.1 x 1.2 mm^3^. The pCASL scan was acquired with Fair QII using a multi-slice interleaved saturation sequence with the following parameters: repetition time (TR) 3840 ms, echo time (TE) 40.7 ms, field of view 240 mm x 240 mm, 54 slices with 2.5 mm thickness, image matrix 96 x 96, and voxel size 2.5 x 2.5 x 2.5 mm^3^. The label duration was 1500ms and post-labeling delay (PLD) was 1800ms. The total acquisition time was 7min 18sec including a M0 image and 7 pairs of label and control images.

For each individual, all pCASL images were aligned to the first pCASL image and averaged. The mean pCASL image was coregistered to the individual’s structural MRI image, and the structural image was coregistered to an averaged brain mask template. Then to obtain gray matter maps, the individual T1-weighted structural MRI images were segmented, and the gray matter images were normalized and smoothed with a 6 mm isotropic full width at half maximum (FWHM) Gaussian kernel.

To create a gray matter mask, the gray matter tissue map from the SPM12-segmented structural MRI was coregistered to the mean perfusion image, thresholded (> 0.75), smoothed (isotropic 6 mm FWHM Gaussian kernel), and binarized (> 0.20). This individualized gray matter mask was then applied to the mean CBF map, which was normalized to Montreal Neurological Institute (MNI) atlas space using nonlinear transformation parameters from the segmentation step, resampled to 2 mm/side isotropic voxels, and smoothed with an isotropic 6 mm FWHM Gaussian kernel, with the cerebellum excluded in order to remove individual variability in this region due to the imaging method.

2.3 A$\beta$ PET

For the [^18^F]florbetapir scans, 10 mCi $\pm10\%$ of [^18^F]florbetapir was injected intravenously, followed by a 50-minute uptake period. For [^18^F]florbetaben scans, 8.1 mCi $\pm10\%$of [^18^F]florbetaben was injected intravenously, followed by a 90-minute uptake period. Participants were then imaged on the same Siemens Biograph mCT PET/CT scanner for 20 minutes with continuous list mode data acquisition. For both types of scans, a computed tomography scan was acquired for scatter and attenuation correction. The list mode data were then binned into four 5-minute frames and reconstructed using parameters defined in the Alzheimer’s Disease Neuroimaging Initiative (ADNI) 2 protocol (Jagust et al., 2015; https://adni.loni.usc.edu/methods/pet-analysis-method/pet-analysis/). These parameters include corrections for scatter and random coincidence events, attenuation, and radionuclide decay. Each 5-minute frame was spatially aligned to the participant’s T1-weighted structural MRI using coreg, normalized to MNI space, and smoothed with an 8 mm FWHM Gaussian kernel, all using SPM12. The spatially aligned PET images were averaged to create a single image from the total acquisition time period for each individual (50-70 minutes for [^18^F]florbetapir and 90-110 minutes for [^18^F]florbetaben), and standardized uptake value ratio (SUVR) images were generated by intensity normalizing with the average radioactivity from a whole cerebellar region of interest from the Centiloid project (http://www.gaain.org/Centiloid-project/).

2.4 Tau PET

10 mCi $\pm10\%$ of [^18^F]flortaucipir was injected intravenously, followed by a 75-minute uptake period. Individuals were then imaged on the same Siemens Biograph mCT PET/CT scanner for 30 minutes with continuous list mode data acquisition. These data were binned into six 5-minute frames, and scans were reconstructed using a standard scanner software program (Siemens, Knoxville, TN) according to the ADNI2 protocol (Jagust et al., 2015; https://adni.loni.usc.edu/methods/pet-analysis-method/pet-analysis/). The 5-minute frames corresponding to 80-100 minutes post-injection were spatially aligned to each participant’s T1 image and normalized to MNI space, averaged to create a single image, intensity normalized by the cerebellar crus (Schwarz et. al., 2016) to create SUVR images, and smoothed with an 8 mm FWHM Gaussian kernel, all using SPM12.

Supplementary Table 1. Correlation of age with lobar CBF in CN.

|  | Independent Variable retained in Model | Standardized Coefficient ($\beta$) | *t* | *p* |
| --- | --- | --- | --- | --- |
| Frontal | Age | $-$0.511 | $-$3.147 | 0.004 |
| Limbic | Age | $-$0.438 | $-$2.580 | 0.015 |
| Parietal | Age | $-$0.512 | $-3$.153 | 0.004 |
| Temporal | Age | $-$0.514 | $-$3.173 | 0.004 |
| Occipital | Age | $-$0.455 | $-$2.707 | 0.011 |

Supplementary Table 2. Correlation of reported hypertension and sex with lobar CBF in MCI.

|  | Independent Variable retained in Model | Standardized Coefficient ($\beta$) | *t* | *p* |
| --- | --- | --- | --- | --- |
| Frontal | Reported Hypertension | $-$0.421 | $-$2.129 | 0.045 |
| Limbic | Sex | 0.472 | 2.456 | 0.023 |
| Temporal | Sex | 0.451 | 2.313 | 0.031 |
| Occipital | Sex | 0.506 | 2.686 | 0.014 |

Supplementary Table 3. Regions of negative association between age and voxel-wise CBF in the full sample. *p*$\leq$0.001; *k*$\geq$50 Peak effects reported in MNI coordinates (mm).

| k | ROI | x | y | z | Brodmann area | *t* | Uncorrected cluster-wise *p* |
| --- | --- | --- | --- | --- | --- | --- | --- |
| 993 | left superior temporal | $-56$ | $12$ | $-6$ | 22 | 4.98 | <0.001 |
|  | left primary auditory | $-48$ | $-16$ | $8$ | 41 | 4.95 |  |
|  | left pre- and supplementary motor | $-58$ | $0$ | 0 | 6 | 4.27 |  |
|  | left primary sensory | $-64$ | $-16$ | 10 | 1 | 3.93 |  |
|  | left primary motor | $-66$ | $-6$ | $14$ | 4 | 3.92 |  |
|  | left temporal pole | $-42$ | $12$ | $-20$ | 38 | 3.84 |  |
|  | left insula | $-42$ | $6$ | $-10$ | 13 | 3.79 |  |
|  | left supramarginal | $-48$ | $-24$ | 12 | 40 | 3.78 |  |
|  | left temporal pole | $-38$ | 16 | $-24$ | 38 | 3.72 |  |
|  | left insula | $-38$ | $-20$ | $-$2 | 13 | 3.64 |  |
|  | left insula | $-42$ | $-2$ | $-8$ | 13 | 3.52 |  |
|  | left insula | $-40$ | $-8$ | $-6$ | 13 | 3.50 |  |
| 543 | right temporal pole | $32$ | 20 | $-42$ | 38 | 4.96 | <0.001 |
|  | right temporal pole | 26 | 12 | $-34$ | 38 | 4.48 |  |
| 748 | right thalamus | $6$ | $-4$ | 4 | N/A | 4.51 | <0.001 |
|  | right caudate | 8 | 6 | 10 | N/A | 4.17 |  |
|  | left thalamus | $-2$ | $-4$ | 0 | N/A | 4.16 |  |
|  | left hippocampus | $-18$ | $-10$ | $-16$ | N/A | 3.80 |  |
|  | left caudate | $-6$ | 8 | 8 | N/A | 3.38 |  |
| 392 | left primary motor | $-18$ | $-28$ | 76 | 4 | 4.74 | 0.001 |
|  | left primary motor | $-44$ | $-18$ | 60 | 4 | 3.80 |  |
| 454 | right pre- and supplementary motor | 38 | $-18$ | 62 | 6 | 4.44 | <0.001 |
|  | right primary motor | 44 | $-18$ | 52 | 4 | 4.22 |  |
|  | right pre- and supplementary motor | 18 | $-24$ | 78 | 6 | 3.76 |  |
|  | right pre- and supplementary motor | 26 | $-18$ | 62 | 6 | 3.73 |  |
| 314 | right primary visual | 18 | $-84$ | $12$ | 17 | 4.43 | 0.002 |
|  | right secondary visual | 10 | $-98$ | 12 | 18 | 3.71 |  |
| 780 | right primary sensory | 46 | $-16$ | 12 | 1 | 4.34 | <0.001 |
|  | right pars orbitalis | 48 | 20 | $-14$ | 47 | 4.18 |  |
|  | right hippocampus | 36 | $-14$ | $-20$ | N/A | 4.04 |  |
|  | right insula | 40 | 2 | $-12$ | 13 | 4.03 |  |
|  | right insula | 42 | $-8$ | $-8$ | 13 | 4.00 |  |
|  | right superior temporal | 64 | $-2$ | 4 | 22 | 3.97 |  |
|  | right hippocampus | 36 | $-20$ | $-12$ | N/A | 3.79 |  |
|  | right primary auditory | 40 | $-18$ | 4 | 41 | 3.61 |  |
|  | right temporal pole | 42 | 20 | $-26$ | 38 | 3.53 |  |
|  | right insula | 40 | $-8$ | 10 | 13 | 3.43 |  |
|  | right hippocampus | 36 | $-28$ | $-10$ | N/A | 3.41 |  |
| 91 | right parahippocampal | 36 | $-16$ | $-36$ | 36 | 3.72 | 0.065 |
|  | right inferior temporal | 34 | $-8$ | $-38$ | 20 | 3.60 |  |
| 83 | left dorsal anterior cingulate | $-4$ | 42 | 2 | 32 | 4.10 | 0.077 |
|  | right dorsal anterior cingulate | 4 | 42 | 4 | 32 | 3.40 |  |
| 55 | left pre- and supplementary motor | $-28$ | $-4$ | 64 | 6 | 4.00 | 0.142 |
| 67 | left primary sensory | $-48$ | $-32$ | 50 | 1 | 3.96 | 0.108 |
|  | left primary sensory | $-42$ | $-36$ | 56 | 1 | 3.76 |  |
| 82 | left temporal pole | $-30$ | $-2$ | $-44$ | 38 | 3.66 | 0.078 |
|  | left parahippocampal | $-30$ | 0 | $-34$ | 36 | 3.63 |  |
| 85 | right pars triangularis | 36 | 28 | 8 | 45 | 3.93 | 0.074 |
|  | right pars opercularis | 40 | 20 | 14 | 44 | 3.62 |  |
| 51 | left primary sensory | $-36$ | $-24$ | 46 | 1 | 3.89 | 0.157 |
|  | left primary sensory | $-40$ | $-20$ | 38 | 1 | 3.61 |  |

ROI: region of interest

Supplementary Table 4. Regions of negative association between age and voxel-wise CBF in CN. *p*$\leq$0.001; *k*$\geq$50. Peak effects reported in MNI coordinates (mm).

| k | ROI | x | y | z | Brodmann area | *t* | Uncorrected cluster-wise *p* |
| --- | --- | --- | --- | --- | --- | --- | --- |
| 136 | right temporal pole | 28 | 14 | $-36$ | 38 | 4.60 | 0.028 |
| 59 | right caudate | 12 | 22 | $-6$ | N/A | 4.23 | 0.129 |

ROI: region of interest

Supplementary Table 5. Regions of negative association between reported hypertension and voxel-wise CBF in the full sample. *p*$\leq$0.001; *k*$\geq$50. Peak effects reported in MNI coordinates (mm).

| k | ROI | x | y | z | Brodmann area | *t* | Uncorrected cluster-wise *p* |
| --- | --- | --- | --- | --- | --- | --- | --- |
| 118 | left temporal pole | $-38$ | $4$ | $-36$ | 38 | 4.87 | 0.037 |
|  | left inferior temporal | $-40$ | $-4$ | $-32$ | 20 | 3.66 |  |
| 175 | right anterior prefrontal cortex | 8 | 46 | $-6$ | 10 | 4.69 | 0.014 |
|  | right dorsal anterior cingulate | 10 | 40 | 12 | 32 | 4.21 |  |
|  | right dorsal anterior cingulate | 6 | 36 | $-4$ | 32 | 3.96 |  |
|  | right dorsal anterior cingulate | 10 | 44 | 4 | 32 | 3.36 |  |
| 153 | right frontal eye fields | 42 | 22 | 48 | 8 | 4.21 | 0.020 |
|  | right frontal eye fields | 28 | 28 | 48 | 8 | 3.91 |  |
| 60 | right frontal eye fields | 36 | 8 | 56 | 8 | 4.13 | 0.122 |
| 161 | left pars orbitalis | $-24$ | 28 | $-12$ | 47 | 4.10 | 0.017 |
|  | left orbital frontal | $-20$ | 32 | $-16$ | 11 | 4.02 |  |
|  | left orbital frontal | $-22$ | 38 | $-12$ | 11 | 3.77 |  |
| 229 | left dorsolateral prefrontal cortex | $-20$ | 46 | 34 | 9 | 4.00 | 0.006 |
|  | left dorsolateral prefrontal cortex | $-12$ | 52 | 34 | 9 | 3.92 |  |
|  | left frontal eye fields | $-12$ | 46 | 40 | 8 | 3.83 |  |
|  | left dorsolateral prefrontal cortex | $-4$ | 50 | 36 | 9 | 3.71 |  |
|  | left dorsolateral prefrontal cortex | $-26$ | 40 | 34 | 9 | 3.56 |  |
|  | left anterior prefrontal cortex | $-28$ | 48 | 28 | 10 | 3.50 |  |
| 57 | left dorsolateral prefrontal cortex | $-46$ | 24 | 34 | 9 | 3.96 | 0.131 |
| 56 | right primary sensory | 32 | $-34$ | 42 | 1 | 3.43 | 0.134 |
| 56 | right pars opercularis | 56 | 10 | 4 | 44 | 3.95 | 0.134 |
| 50 | right pars orbitalis | 48 | 22 | $-2$ | 47 | 3.79 | 0.156 |
| 51 | right pars orbitalis | 50 | 38 | $-14$ | 47 | 3.72 | 0.152 |
| 50 | left pre- and supplementary motor | $-32$ | $-6$ | 60 | 6 | 3.70 | 0.156 |
|  | left pre- and supplementary motor | $-38$ | 0 | 56 | 6 | 3.57 |  |
| 66 | right ventral anterior cingulate | 2 | $-20$ | 40 | 24 | 3.65 | 0.107 |

ROI: region of interest

Supplementary Table 6. Regions of negative association between reported hypertension and voxel-wise CBF in MCI. *p*$\leq$0.001; *k*$\geq$50. Peak effects reported in MNI coordinates (mm).

| k | ROI | x | y | z | Brodmann area | *t* | Uncorrected cluster-wise *p* |
| --- | --- | --- | --- | --- | --- | --- | --- |
| 176 | right pars orbitalis | $34$ | $34$ | $-16$ | 47 | 5.01 | 0.003 |
|  | right pars orbitalis | 56 | 36 | $-2$ | 47 | 5.00 |  |
|  | right pars orbitalis | 52 | 38 | $-10$ | 47 | 4.55 |  |
|  | right pars orbitalis | 30 | 30 | $-8$ | 47 | 4.22 |  |

ROI: region of interest

Supplementary Table 7. Regions of negative association between reported hypertension and voxel-wise CBF with the vascular risk score included in the full sample. *p*$\leq$0.001; *k*$\geq$50. Peak effects reported in MNI coordinates (mm).

| k | ROI | x | y | z | Brodmann area | *t* | Uncorrected cluster-wise *p* |
| --- | --- | --- | --- | --- | --- | --- | --- |
| 178 | left dorsal anterior cingulate | $-8$ | $30$ | $-8$ | 32 | 3.89 | 0.013 |

ROI: region of interest

Supplementary Table 7. Regions of negative association between reported hypertension and voxel-wise CBF with the vascular risk score included in MCI. *p*$\leq$0.001; *k*$\geq$50. Peak effects reported in MNI coordinates (mm).

| k | ROI | x | y | z | Brodmann area | *t* | Uncorrected cluster-wise *p* |
| --- | --- | --- | --- | --- | --- | --- | --- |
| 119 | right orbital frontal | 16 | 42 | $-24$ | 11 | 4.28 | 0.010 |
| 1398 | right secondary visual | 14 | $-100$ | 12 | 18 | 9.30 | <0.001 |
|  | right secondary visual | 18 | $-88$ | 16 | 18 | 5.19 |  |
|  | right primary visual | 18 | $-98$ | 2 | 17 | 5.05 |  |
|  | right secondary visual | 20 | $-94$ | 8 | 18 | 4.83 |  |
|  | right secondary visual | 8 | $-74$ | $-8$ | 18 | 4.73 |  |
|  | right primary visual | 0 | $-96$ | 10 | 17 | 4.41 |  |
|  | right secondary visual | 34 | $-94$ | $-4$ | 18 | 4.28 |  |
|  | right secondary visual | 26 | $-98$ | $-14$ | 18 | 4.26 |  |
|  | right secondary visual | 28 | $-94$ | 14 | 18 | 4.00 |  |
|  | right secondary visual | 32 | $-88$ | 10 | 18 | 3.84 |  |
| 81 | right medial temporal | 62 | $-20$ | $-20$ | 21 | 4.82 | 0.029 |
| 269 | right anterior prefrontal cortex | 6 | 46 | $-10$ | 10 | 7.35 | <0.001 |
|  | left anterior prefrontal cortex | $-10$ | 44 | $-8$ | 10 | 6.85 |  |
|  | right orbital frontal | 10 | 54 | $-14$ | 11 | 5.12 |  |
|  | left orbital frontal | $-10$ | 50 | $-16$ | 11 | 5.03 |  |
|  | right dorsal anterior cingulate | 6 | 38 | $-2$ | 32 | 3.90 |  |
| 142 | left pre- and supplementary motor | $-10$ | $-14$ | 78 | 6 | 4.51 | 0.006 |
|  | left pre- and supplementary motor | $-20$ | $-10$ | 74 | 6 | 4.44 |  |
| 65 | left pre- and supplementary motor | $-22$ | 24 | 60 | 6 | 4.04 | 0.047 |
| 71 | left thalamus | $-8$ | $-14$ | $-2$ | N/A | 5.01 | 0.039 |

ROI: region of interest

Supplementary Table 7. Regions of positive association between female sex and voxel-wise CBF. *p*$\leq$0.001; *k*$\geq$50. Peak effects reported in MNI coordinates (mm).

| k | ROI | x | y | z | Brodmann area | *t* | Uncorrected cluster-wise *p* |
| --- | --- | --- | --- | --- | --- | --- | --- |
| 784 | right temporal pole | $62$ | $0$ | $-14$ | 38 | 5.84 | <0.001 |
|  | right medial temporal | $62$ | $-$2 | $-24$ | 21 | 4.97 |  |
|  | right medial temporal | 70 | $-24$ | $-16$ | 21 | 4.44 |  |
|  | right medial temporal | 68 | $-32$ | $-18$ | 21 | 4.39 |  |
|  | right inferior temporal | 56 | 2 | $-36$ | 20 | 3.81 |  |
| 1362 | right dorsolateral prefrontal cortex | 0 | 46 | 22 | 9 | 4.20 | <0.001 |
|  | right orbital frontal | 8 | 56 | $-26$ | 11 | 4.16 |  |
|  | right dorsal anterior cingulate | 0 | 48 | 10 | 32 | 4.15 |  |
|  | right anterior prefrontal cortex | 12 | 68 | $-10$ | 10 | 4.01 |  |
|  | right anterior prefrontal cortex | 4 | 58 | 14 | 10 | 3.58 |  |
|  | right anterior prefrontal cortex | 22 | 54 | -18 | 10 | 3.49 |  |
| 844 | left medial temporal | $-66$ | $-12$ | $-6$ | 21 | 5.10 | <0.001 |
|  | left medial temporal | $-64$ | $-10$ | $-26$ | 21 | 5.07 |  |
|  | left temporal pole | $-60$ | 6 | $-12$ | 38 | 4.38 |  |
|  | left pars triangularis | $-52$ | 16 | 2 | 45 | 3.30 |  |
| 403 | left orbital frontal | $-24$ | 52 | $-18$ | 11 | 4.74 | 0.001 |
|  | left pars orbitalis | $-44$ | 40 | $-18$ | 47 | 4.12 |  |
|  | left anterior prefrontal cortex | $-42$ | 52 | $-10$ | 10 | 3.79 |  |
|  | left anterior prefrontal cortex | $-36$ | 50 | $-2$ | 10 | 3.47 |  |
|  | left pars orbitalis | $-46$ | 36 | $-20$ | 47 | 3.29 |  |
| 955 | right ventral posterior cingulate | 22 | $-64$ | 8 | 23 | 4.67 | <0.001 |
|  | right primary visual | 14 | $-62$ | 10 | 17 | 4.44 |  |
|  | right primary visual | 22 | $-80$ | 10 | 17 | 4.19 |  |
|  | right visual association | 44 | $-86$ | 12 | 19 | 4.18 |  |
|  | right secondary visual | 30 | $-88$ | 14 | 18 | 4.10 |  |
|  | right secondary visual | 16 | $-46$ | 2 | 18 | 3.81 |  |
|  | right visual association | 22 | $-48$ | $-4$ | 19 | 3.70 |  |
|  | right dorsal posterior cingulate | 4 | $-68$ | 20 | 31 | 3.53 |  |
|  | right dorsal posterior cingulate | 10 | $-66$ | 22 | 31 | 3.53 |  |
| 251 | right visual association | 52 | $-76$ | $-6$ | 19 | 4.47 | 0.005 |
|  | right fusiform | 48 | $-66$ | 2 | 37 | 4.16 |  |
|  | right visual association | 48 | $-72$ | 10 | 19 | 3.84 |  |
| 81 | left medial temporal | $-68$ | $-38$ | $-4$ | 21 | 4.40 | 0.080 |
| 61 | left parahippocampal | $-34$ | $-20$ | $-30$ | 36 | 4.34 | 0.124 |
| 155 | right anterior prefrontal cortex | 42 | 48 | $-18$ | 10 | 4.26 | 0.020 |
|  | right pars orbitalis | 50 | 44 | $-10$ | 47 | 3.62 |  |
| 159 | left secondary visual | $-34$ | $-94$ | 8 | 18 | 3.88 | 0.019 |
| 109 | right secondary visual | 22 | $-94$ | $-16$ | 18 | 3.36 | 0.046 |
| 121 | right insula | 44 | 14 | $-4$ | 13 | 3.69 | 0.037 |
| 51 | left insula | $-44$ | $-4$ | 2 | 13 | 3.95 | 0.157 |
| 139 | left secondary visual | $-20$ | $-96$ | $-14$ | 18 | 3.89 | 0.027 |
|  | left secondary visual | $-24$ | $-90$ | $-20$ | 18 | 3.57 |  |
| 70 | right secondary visual | 2 | $-70$ | 0 | 18 | 3.88 | 0.101 |
| 86 | left pars orbitalis | $-40$ | 22 | $-8$ | 47 | 3.86 | 0.072 |
|  | left temporal pole | $-40$ | 22 | $-22$ | 38 | 3.38 |  |
| 136 | right temporal pole | 40 | 24 | $-34$ | 38 | 3.85 | 0.028 |
|  | right temporal pole | 44 | 22 | $-30$ | 38 | 3.84 |  |
| 88 | right parahippocampal | 28 | $-24$ | $-26$ | 36 | 3.84 | 0.069 |
| 55 | right temporal pole | 32 | 4 | $-36$ | 38 | 3.79 | 0.142 |
| 77 | right secondary visual | 10 | $-88$ | 24 | 18 | 3.71 | 0.087 |
| 75 | right superior temporal | 44 | $-42$ | 6 | 22 | 3.70 | 0.091 |
| 56 | right insula | 32 | $-22$ | 18 | 13 | 3.59 | 0.139 |

ROI: region of interest

Supplementary Figure 1. Negative correlations between voxel-wise CBF and age. Covariates are sex, diagnostic group and total gray matter volume. Images in left column illustrated at *p*$\leq$0.001; *k*$\geq$50. Right column shows $\beta$ maps.

| Full Sample | 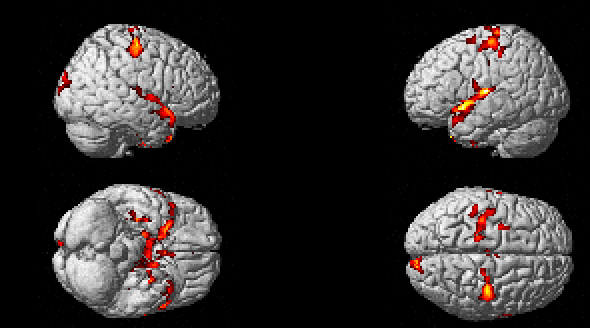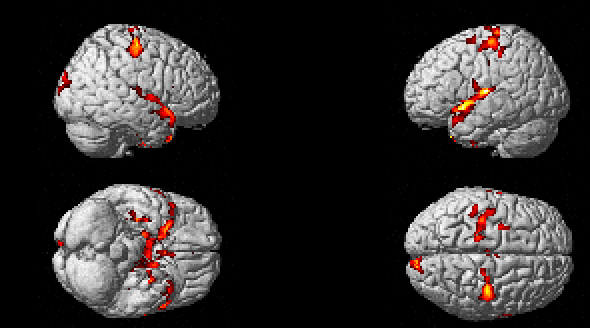 | 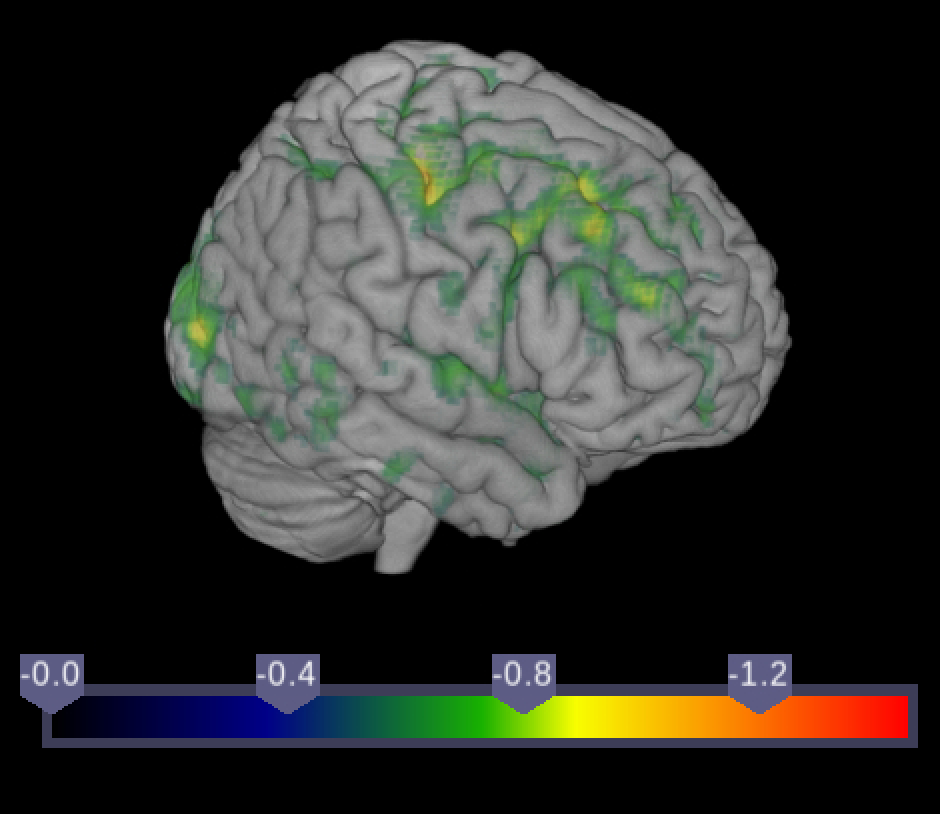 |
| --- | --- | --- |
| CN | 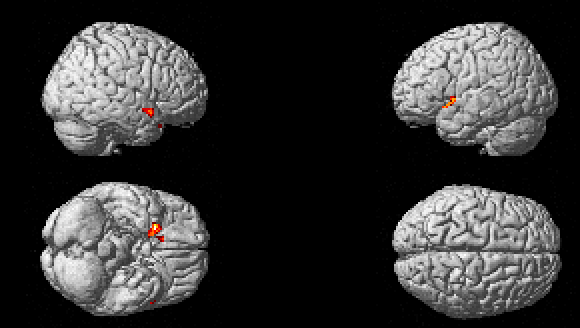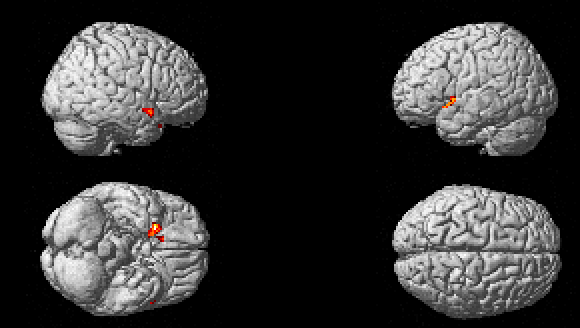 | 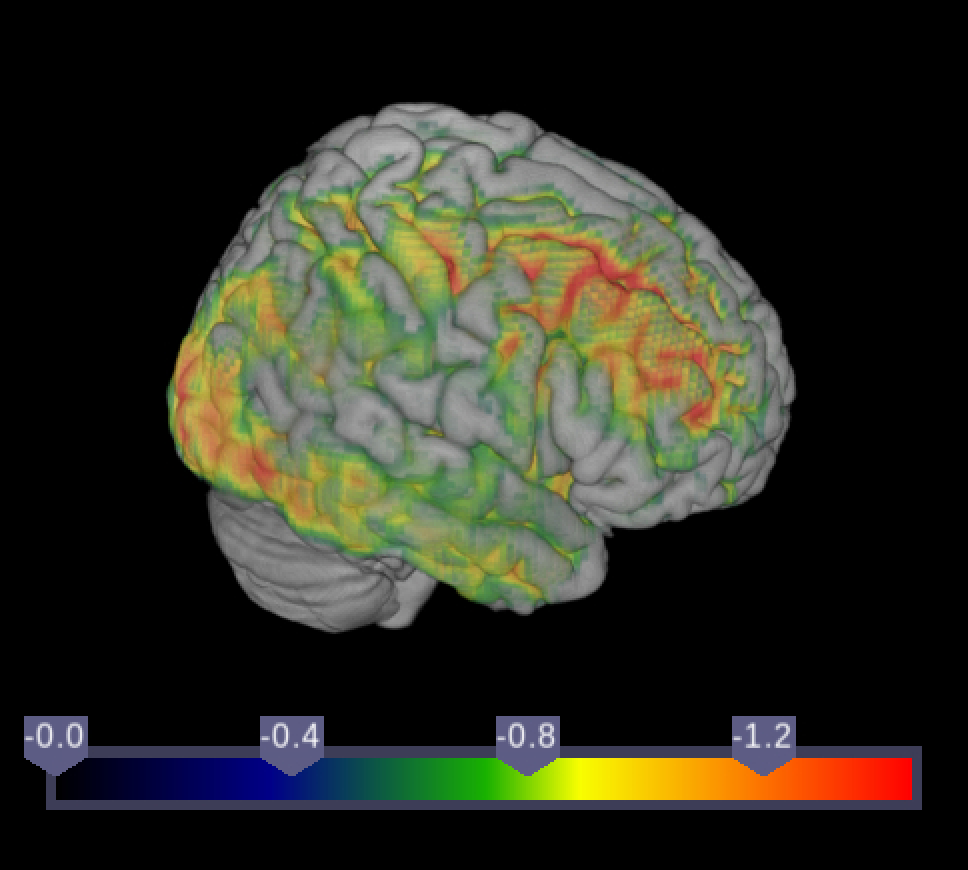 |

Supplementary Figure 2. Positive correlations between voxel-wise CBF and female sex. Covariates are age, diagnostic group and total gray matter volume. Images in left column illustrated at *p*$\leq$0.001; *k*$\geq$50. Right column shows $\beta$ maps.

| Full Sample | 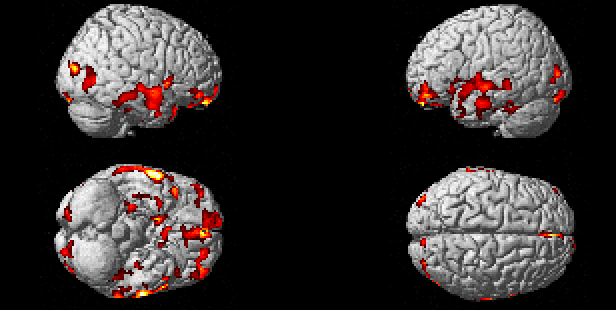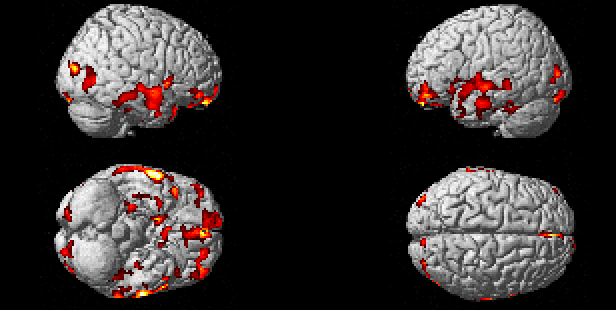 | 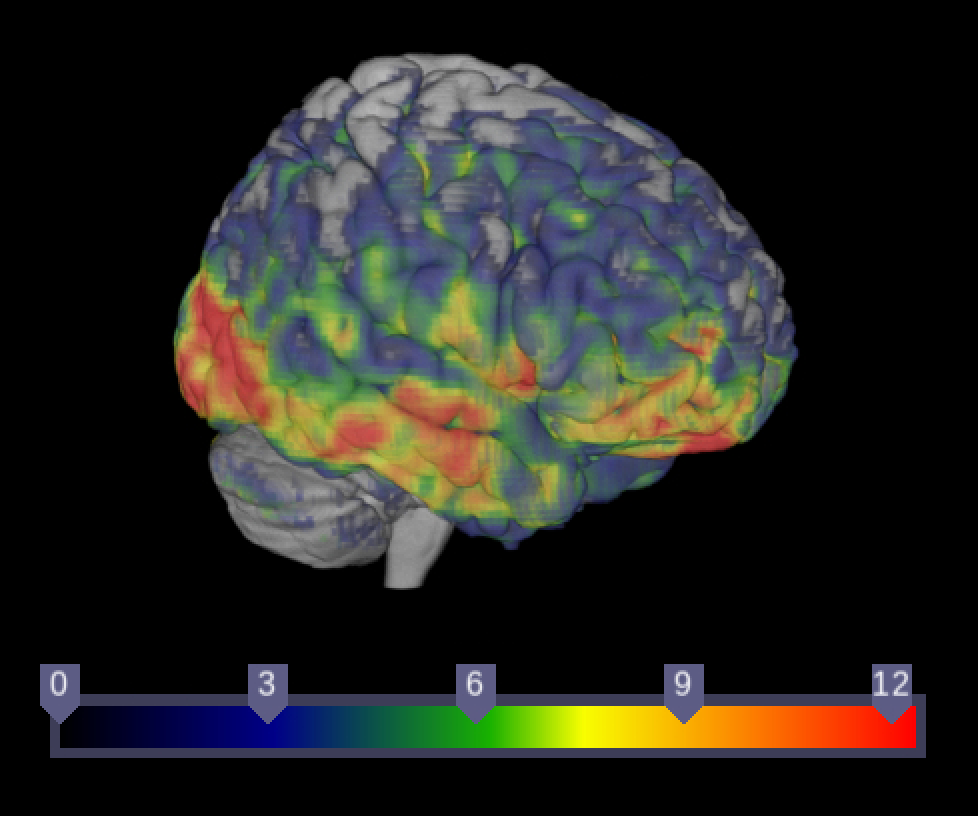 |
| --- | --- | --- |
| MCI | *no significant clusters* | 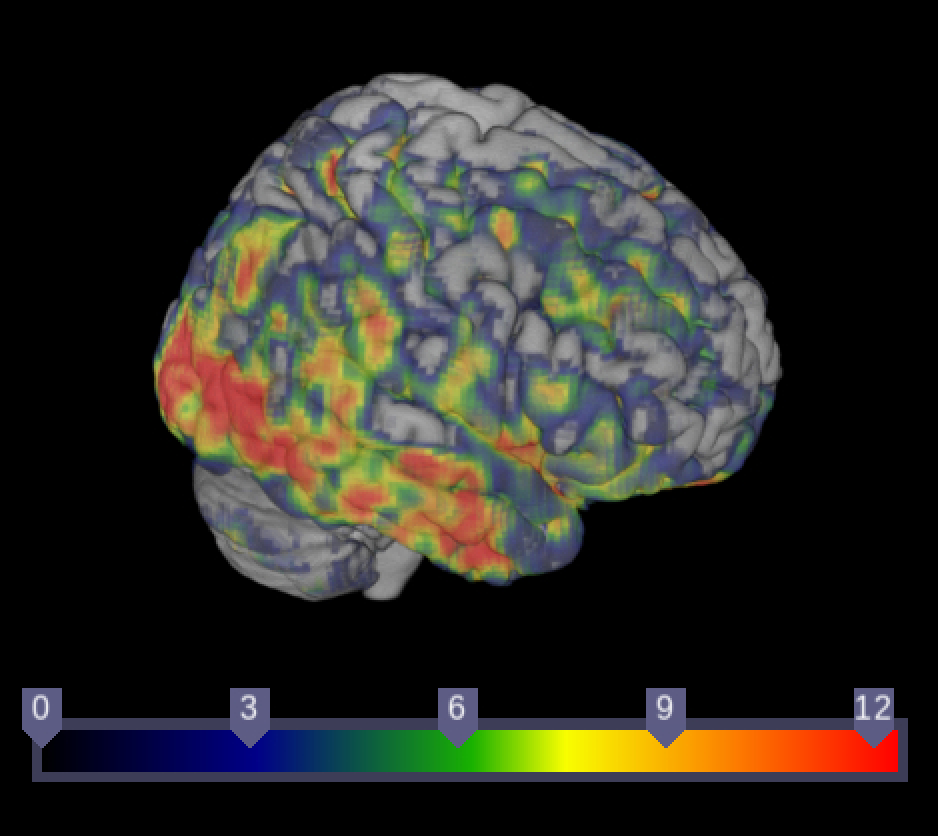 |

Supplementary Table 8. Regions of negative association between MTL tau SUVR and voxel-wise CBF in the full sample. *p*$\leq$0.001; *k*$\geq$50. Peak effects reported in MNI coordinates (mm).

| k | ROI | x | y | z | Brodmann area | *t* | Uncorrected cluster-wise *p* |
| --- | --- | --- | --- | --- | --- | --- | --- |
| 179 | left parahippocampal | $-22$ | $-20$ | $-20$ | 36 | 5.14 | 0.014 |
|  | left parahippocampal | $-18$ | $-16$ | $-26$ | 36 | 4.31 |  |

Supplementary Table 9. Regions of negative association between MTL tau SUVR and voxel-wise CBF in SCD. *p*$\leq$0.001; *k*$\geq$50. Peak effects reported in MNI coordinates (mm).

| k | ROI | x | y | z | Brodmann area | *t* | Uncorrected cluster-wise *p* |
| --- | --- | --- | --- | --- | --- | --- | --- |
| 146 | left primary sensory | $-22$ | $-30$ | $74$ | 1 | 5.59 | 0.009 |
| 157 | right primary motor | 26 | $-28$ | 74 | 4 | 4.95 | 0.007 |
| 84 | right visual motor | 34 | $-54$ | 62 | 7 | 4.78 | 0.039 |
|  | right visual motor | 34 | $-42$ | 58 | 7 | 3.90 |  |
| 118 | right primary sensory | 60 | $-14$ | 44 | 1 | 4.04 | 0.017 |

Supplementary Table 10. Regions of positive association between the MTL tau SUVR and reported hypertension interaction variable and voxel-wise CBF in SCD. *p*$\leq$0.001; *k*$\geq$50. Peak effects reported in MNI coordinates (mm).

| k | ROI | x | y | z | Brodmann area | *t* | Uncorrected cluster-wise *p* |
| --- | --- | --- | --- | --- | --- | --- | --- |
| 114 | right superior temporal | 44 | $-8$ | $-14$ | 22 | 5.59 | 0.017 |
| 95 | right fusiform | 44 | $-52$ | 12 | 37 | 6.22 | 0.027 |
|  | right angular gyrus | 50 | $-52$ | 22 | 39 | 3.71 |  |
| 66 | right primary motor | 46 | $-18$ | 56 | 4 | 6.10 | 0.059 |
|  | right primary sensory | 54 | $-20$ | 54 | 1 | 5.11 |  |
| 109 | left pre- and supplementary motor | $-14$ | $-22$ | 44 | 6 | 5.99 | 0.019 |

Supplementary Table 11. Regions of negative association between the MTL tau SUVR and *APOE* ε4 status interaction variable and voxel-wise CBF in SCD. *p*$\leq$0.001; *k*$\geq$50. Peak effects reported in MNI coordinates (mm).

| k | ROI | x | y | z | Brodmann area | *t* | Uncorrected cluster-wise *p* |
| --- | --- | --- | --- | --- | --- | --- | --- |
| 173 | right pre- and supplementary motor | 24 | $2$ | $50$ | 6 | 7.74 | 0.003 |
|  | right pre- and supplementary motor | 38 | $-6$ | 46 | 6 | 4.84 |  |
| 153 | right angular | 48 | $-48$ | 38 | 39 | 7.21 | 0.004 |
|  | right angular | 48 | $-48$ | 28 | 39 | 4.94 |  |
|  | right visual motor | 46 | $-44$ | 50 | 7 | 4.57 |  |
| 501 | left primary sensory | $-44$ | $-22$ | 40 | 1 | 6.88 | <0.001 |
|  | left sensory association | $-10$ | $-36$ | 58 | 5 | 5.53 |  |
|  | left visual motor | $-30$ | $-48$ | 56 | 7 | 5.11 |  |
|  | left pre- and supplementary motor | $-56$ | $-8$ | 26 | 6 | 4.88 |  |
|  | left primary motor | $-48$ | $-14$ | 36 | 4 | 4.85 |  |
|  | left pre- and supplementary motor | $-56$ | $-6$ | 30 | 6 | 4.82 |  |
|  | left primary sensory | $-60$ | $-10$ | 20 | 1 | 4.81 |  |
|  | left pre- and supplementary motor | $-54$ | $-4$ | 36 | 6 | 4.79 |  |
|  | left supramarginal | $-40$ | $-40$ | 42 | 40 | 4.67 |  |
|  | left primary motor | $-50$ | $-10$ | 38 | 4 | 4.61 |  |
|  | left primary sensory | $-8$ | $-30$ | 60 | 1 | 4.54 |  |
|  | left supramarginal | $-36$ | $-38$ | 46 | 40 | 4.13 |  |
|  | left visual motor | $-32$ | $-42$ | 46 | 7 | 3.84 |  |
| 258 | right pars orbitalis | 42 | 38 | $-12$ | 47 | 5.19 | <0.001 |
|  | right pars orbitalis | 36 | 38 | $-10$ | 47 | 4.95 |  |
|  | right pars orbitalis | 30 | 32 | $-14$ | 47 | 4.60 |  |
|  | right orbital frontal | 22 | 28 | $-20$ | 11 | 4.31 |  |
|  | right pars orbitalis | 24 | 30 | $-16$ | 47 | 3.94 |  |
| 1173 | right primary motor | 50 | $-14$ | 36 | 4 | 6.73 | <0.001 |
|  | right supramarginal | 54 | $-34$ | 34 | 40 | 6.20 |  |
|  | right superior temporal | 60 | $-40$ | 18 | 22 | 5.69 |  |
|  | right supramarginal | 62 | $-32$ | 30 | 40 | 5.67 |  |
|  | right superior temporal | 60 | $-42$ | 8 | 22 | 5.48 |  |
|  | right supramarginal | 46 | $-32$ | 42 | 40 | 5.45 |  |
|  | right supramarginal | 56 | $-30$ | 44 | 40 | 5.03 |  |
|  | right supramarginal | 38 | $-36$ | 46 | 40 | 5.03 |  |
|  | right primary sensory | 56 | $-22$ | 42 | 1 | 4.92 |  |
|  | right visual motor | 22 | $-46$ | 56 | 7 | 4.91 |  |
|  | right primary sensory | 56 | $-22$ | 30 | 1 | 4.88 |  |
|  | right supramarginal | 50 | $-18$ | 18 | 40 | 4.74 |  |
|  | right primary motor | 52 | $-6$ | 12 | 4 | 4.71 |  |
|  | right primary sensory | 42 | $-32$ | 56 | 1 | 4.60 |  |
| 250 | right inferior temporal | 42 | $-24$ | $-18$ | 20 | 6.48 | <0.001 |
|  | right superior temporal | 52 | $-20$ | $-6$ | 22 | 5.86 |  |
|  | right superior temporal | 58 | $-18$ | $-4$ | 22 | 4.64 |  |
|  | right medial temporal | 60 | $-16$ | $-12$ | 21 | 4.08 |  |
|  | right caudate | 32 | $-34$ | $-2$ | N/A | 3.88 |  |
|  | right putamen | 32 | $-16$ | 2 | N/A | 3.86 |  |
| 64 | left supramarginal | $-42$ | $-28$ | 20 | 40 | 6.17 | 0.046 |
| 143 | left insula | $-32$ | 6 | 14 | 13 | 4.75 | 0.005 |
| 85 | right pre- and supplementary motor | 52 | 2 | 22 | 6 | 6.05 | 0.024 |
|  | right pre- and supplementary motor | 54 | 6 | 12 | 6 | 3.94 |  |
|  | right pars opercularis | 50 | 8 | 8 | 44 | 3.79 |  |
| 104 | right inferior temporal | 56 | $-16$ | $-28$ | 20 | 6.03 | 0.014 |
|  | right inferior temporal | 54 | $-16$ | $-32$ | 20 | 5.61 |  |
|  | right medial temporal | 60 | $-16$ | $-20$ | 21 | 4.68 |  |
| 205 | left dorsal posterior cingulate | $-4$ | $-52$ | 40 | 31 | 5.78 | 0.001 |
|  | left dorsal posterior cingulate | $-6$ | $-54$ | 34 | 31 | 5.48 |  |
| 226 | right pars opercularis | 36 | 18 | 18 | 44 | 5.78 | 0.001 |
|  | right dorsolateral prefrontal cortex | 42 | 20 | 24 | 9 | 4.86 |  |
|  | right insula | 32 | 8 | 10 | 13 | 4.64 |  |
|  | right pars opercularis | 34 | 14 | 10 | 44 | 4.21 |  |
| 50 | left dorsolateral prefrontal cortex | $-40$ | 46 | 6 | 46 | 5.56 | 0.073 |
| 77 | left hypothalamus | $-2$ | $-2$ | $-6$ | N/A | 3.99 | 0.031 |
|  | right hypothalamus | 2 | $-6$ | $-10$ | N/A | 3.83 |  |
| 85 | left visual motor | $-14$ | $-44$ | 54 | 7 | 3.79 | 0.024 |
| 120 | left pars orbitalis | $-32$ | 36 | $-8$ | 47 | 5.31 | 0.009 |
| 53 | left pre- and supplementary motor | $-46$ | $-4$ | 12 | 6 | 5.31 | 0.066 |
|  | left pre- and supplementary motor | $-46$ | 2 | 10 | 6 | 5.02 |  |
| 112 | left orbital frontal | $-10$ | 24 | $-16$ | 11 | 5.23 | 0.011 |
|  | left dorsal anterior cingulate | $-4$ | 22 | $-8$ | 32 | 4.80 |  |
|  | left orbital frontal | $-16$ | 18 | $-18$ | 11 | 3.99 |  |
| 72 | right fusiform | 54 | $-54$ | $-22$ | 37 | 5.12 | 0.036 |
|  | right fusiform | 54 | $-56$ | $-12$ | 37 | 4.36 |  |
| 103 | left pre- and supplementary motor | $-24$ | $-4$ | 50 | 6 | 5.03 | 0.014 |
| 119 | left angular | $-50$ | $-42$ | 34 | 39 | 4.96 | 0.009 |
|  | left angular | $-46$ | $-42$ | 28 | 39 | 4.66 |  |
|  | left supramarginal | $-54$ | $-36$ | 38 | 40 | 4.31 |  |
|  | left supramarginal | $-48$ | $-32$ | 38 | 40 | 3.91 |  |
| 67 | right anterior prefrontal cortex | 16 | 60 | $-10$ | 10 | 4.84 | 0.042 |
|  | right anterior prefrontal cortex | 20 | 58 | $-2$ | 10 | 4.28 |  |
| 157 | left fusiform | $-54$ | $-54$ | $-16$ | 37 | 4.77 | 0.004 |
|  | left fusiform | $-60$ | $-54$ | $-6$ | 37 | 4.37 |  |
|  | left angular | $-58$ | $-56$ | 6 | 39 | 4.36 |  |
|  | left fusiform | $-58$ | $-60$ | $-2$ | 37 | 4.22 |  |
|  | left fusiform | $-66$ | $-52$ | $-12$ | 37 | 3.87 |  |
|  | left visual association | $-54$ | $-66$ | 2 | 19 | 3.87 |  |
|  | left angular | $-58$ | $-52$ | 14 | 39 | 3.79 |  |
| 137 | right thalamus | 10 | $-20$ | 10 | N/A | 4.18 | 0.006 |

Supplementary Table 12. Regions of positive association between the MTL tau SUVR and *APOE* ε4 status interaction variable and voxel-wise CBF in MCI. *p*$\leq$0.001; *k*$\geq$50. Peak effects reported in MNI coordinates (mm).

| k | ROI | x | y | z | Brodmann area | *t* | Uncorrected cluster-wise *p* |
| --- | --- | --- | --- | --- | --- | --- | --- |
| 111 | right fusiform | 66 | $-44$ | $-20$ | 37 | 4.20 | 0.014 |

Supplementary Table 13. Regions of positive association between global cortical Centiloid and voxel-wise CBF in CN. p<0.001; k>50. Peak effects reported in MNI coordinates (mm).

| k | ROI | x | y | z | Brodmann area | *t* | Uncorrected cluster-wise *p* |
| --- | --- | --- | --- | --- | --- | --- | --- |
| 84 | right visual association | 26 | $-92$ | 32 | 19 | 4.36 | 0.075 |

Supplementary Table 14. Regions of negative association between global cortical Centiloid and voxel-wise CBF in MCI. *p*$\leq$0.001; *k*$\geq$50. Peak effects reported in MNI coordinates (mm).

| k | ROI | x | y | z | Brodmann area | *t* | Uncorrected cluster-wise *p* |
| --- | --- | --- | --- | --- | --- | --- | --- |
| 72 | right fusiform | 30 | $-26$ | $-26$ | 37 | 5.59 | 0.046 |

Supplementary Table 15. Regions of positive association between the global cortical Centiloid and reported hypertension interaction variable and voxel-wise CBF in the full sample. *p*$\leq$0.001; *k*$\geq$50. Peak effects reported in MNI coordinates (mm).

| k | ROI | x | y | z | Brodmann area | *t* | Uncorrected cluster-wise *p* |
| --- | --- | --- | --- | --- | --- | --- | --- |
| 79 | left visual motor | $-26$ | $-54$ | 40 | 7 | 3.35 | 0.079 |

Supplementary Table 16. Regions of positive association between the global cortical Centiloid and reported hypertension interaction variable and voxel-wise CBF in SCD. *p*$\leq$0.001; *k*$\geq$50. Peak effects reported in MNI coordinates (mm).

| k | ROI | x | y | z | Brodmann area | *t* | Uncorrected cluster-wise *p* |
| --- | --- | --- | --- | --- | --- | --- | --- |
| 67 | right hippocampus | $34$ | $-18$ | $-12$ | N/A | 5.68 | 0.062 |
|  | right hippocampus | 32 | $-20$ | $-20$ | N/A | 4.44 |  |

Supplementary Table 17. Regions of positive association between the global cortical Centiloid and reported hypertension interaction variable and voxel-wise CBF in MCI. *p*$\leq$0.001; *k*$\geq$50. Peak effects reported in MNI coordinates (mm).

| k | ROI | x | y | z | Brodmann area | *t* | Uncorrected cluster-wise *p* |
| --- | --- | --- | --- | --- | --- | --- | --- |
| 56 | left frontal eye fields | $-34$ | $16$ | $30$ | 8 | 5.22 | 0.063 |
